# Plasma biomarkers distinguish Boston Criteria 2.0 cerebral amyloid angiopathy from healthy controls

**DOI:** 10.1101/2024.09.04.24313038

**Authors:** Ryan T. Muir, Sophie Stukas, Jennifer G. Cooper, Andrew E. Beaudin, Cheryl R. McCreary, Myrlene Gee, Krista Nelles, Nikita Nukala, Janina Valencia, Kristopher M Kirmess, Sandra E. Black, Michael D. Hill, Richard Camicioli, Cheryl L. Wellington, Eric. E Smith

## Abstract

**INTRODUCTION:** Cerebral Amyloid Angiopathy (CAA) is characterized by the deposition of beta-amyloid (Aβ) in small vessels leading to hemorrhagic stroke and dementia. This study examined whether plasma Aβ_42/40,_ phosphorylated-tau (p-tau), neurofilament light chain (NfL) and Glial Fibrillary Acidic Protein (GFAP) differ in CAA and their potential to discriminate Boston Criteria 2.0 probable CAA from healthy controls.

**METHODS:** Plasma Aβ_42/40,_ p-tau-181, NfL and GFAP were quantified using Simoa and Aβ_42/40_ was also independently quantified using immunoprecipitation liquid chromatography mass-spectrometry (IPMS).

**RESULTS:** 45 participants with CAA and 47 healthy controls had available plasma. Aβ_42/40_ ratios were significantly lower in CAA than healthy controls. While p-tau-181 and NfL were elevated in CAA, GFAP was similar. A combination of Aβ_42/40_ (Simoa), p-tau-181, and NfL resulted in an AUC of 0.90 (95% CI: 0.80, 0.95).

**DISCUSSION:** Plasma Aβ_42/40_, p-tau-181 and NfL differ in those with CAA and together can discriminate CAA from healthy controls.

## INTRODUCTION

Spontaneous intracerebral hemorrhage (ICH) confers high morbidity and mortality and may be the harbinger of an underlying neurodegenerative and cerebrovascular disorder: cerebral amyloid angiopathy (CAA).^1-4^ Accurately diagnosing CAA is important as it identifies those with elevated risk of ICH recurrence and dementia.^5-8^ Further, in an era of emerging immunotherapies targeting beta-amyloid in Alzheimer’s Disease (AD), it is imperative to identify those with CAA given the risk of amyloid related imaging abnormalities (ARIA).^9-13^

CAA is characterized by the progressive deposition of beta-amyloid within the media and adventitia of cortical and leptomeningeal small vessels.^14, 15^ On the other hand, AD is typified by extracellular, intraparenchymal, beta-amyloid plaques and intracellular neurofibrillary tangles of hyperphosphorylated tau.^16^ Up to 80% of patients with AD have CAA on pathology.^7, 16^

Currently, the Boston Criteria 2.0, which integrate core clinical and magnetic resonance imaging (MRI) criteria inform a diagnosis of *probable* and *possible* CAA.^4^ However, limitations of these diagnostic criteria include: the requirement of MRI for full sensitivity; hemorrhagic manifestations occur at the latest stage of CAA; CAA cannot be discerned in mixed states where neuroimaging features of hypertensive arteriopathy coincide with those of CAA; and the criteria may have limited diagnostic discriminability in some settings.^4, 15, 17, 18^

Plasma biomarkers are less invasive, accessible, and scalable evaluations of candidate pathophysiologic substrates of CAA and thus could potentially aid in earlier disease detection. Plasma beta-amyloid (the Aβ_42/40_ ratio) and phosphorylated tau (p-tau) herald the presence of AD in reference to cerebrospinal fluid (CSF) and positron emission tomography (PET) signatures.^19, 20^ Furthermore, neurofilament light chain (NfL), an indicator of axonal injury, is elevated in AD and other neurodegenerative states, and emerging evidence suggests glial fibrillary acidic protein (GFAP), a marker of astrocyte activation, is also elevated in AD.^19^ Plasma biomarkers in CAA are under-explored.^19^ In CSF, those with CAA have reduced Aβ_40_ compared to both controls and AD; reduced Aβ_42_ compared to controls (though similar Aβ_42_ compared to AD); reduced Aβ_42/40_ compared to controls (though higher compared to AD); and elevated total-tau and phosphorylated-tau compared to controls, but lower than AD.^21^ The current study evaluated (i) whether plasma Aβ_42_, Aβ_40,_ Aβ_42/40_ ratio, p-tau-181, GFAP and NfL differ in CAA compared to controls and (ii) evaluated biomarker discriminative performance individually and in combination.

## METHODOLOGY

### Research Ethics

The Functional Assessment of Vascular Reactivity in Small Vessel Disease-II (FAVR-II) Study is a prospective cohort study approved by the University of Calgary and University of Alberta Research Ethics Boards (REB). Written informed consent was provided by all participants. The current cross-sectional analysis includes FAVR-II baseline data.

### Study Population

Participants ≥ 55 years of age were recruited between 2015 and 2023 from stroke prevention or cognitive clinics. Healthy controls were community volunteers between the ages of 60 – 85 years old without stroke or cognitive impairment ascertained through medical history and neuropsychological evaluation.^22^ Additionally, participants with substantive alcohol or drug abuse affecting cognition; Montreal Cognitive Assessment (MoCA) scores <13; brain diseases (leukoencephalopathy, multiple sclerosis, neoplasms, Parkinson’s Disease, and other neurologic disorders) were not eligible. Those with CAA presented either with ICH ≥ 90 days before enrollment, transient focal neurologic episodes (TFNEs), or cognitive decline.^22^ A diagnosis of probable CAA was ascertained through Magnetic Resonance Imaging (MRI). A 3.0T MRI (Calgary: Discovery MR750, GE Healthcare, Waukesha, USA; Edmonton: Prisma, Siemens Healthineers, Forchheim, Germany) was obtained including T1-weighted, T2-weighted, proton-density-weighted-T2, fluid attenuated inversion recovery (FLAIR) and T2*-weighted gradient recalled echo (T2*GRE) images. An experienced neurologist (EES) evaluated images for criterion of Boston Criteria 2.0.^4^ A 6 point ordinal CAA small vessel disease score was computed.^23^

### Plasma Collection

At baseline, two 4 mL plastic EDTA tubes of blood were collected. The tubes were mixed by gently inverting 8-10 times then centrifuged at 1200g for 15 minutes. Subsequently, 1.5 mL of plasma from each tube was aliquoted into 0.5 mL polypropylene cryovials (stored at -80°C until analyzed).

### Plasma Biomarker Analysis

Analyses to quantify all plasma biomarker concentrations (pg/mL) were conducted blinded to participant status. Plasma Aβ_42_ and Aβ_40_ concentrations were measured independently by two labs using different methods: (i) liquid chromatography mass-spectrometry (IPMS) analytical platforms previously described and validated at C_2_N Diagnostics (Saint Louis, MO, USA)^24-28^ and (ii) single molecule array (Simoa) at the University of British Columbia. Second, we analyzed plasma Aβ_42_ and Aβ_40_ concentrations with Simoa utilizing the Quanterix Ltd Neurology-4-Plex-E (N4PE) Advantage Kit (cat ID 103670, lot 503768), a multiplex assay that quantifies Aβ_40_, Aβ_42_ NfL and GFAP. Plasma p-tau-181 was measured using version 2.1 Advantage Kit (cat ID 104111, lot 503843). Simoa assays were performed using an HD-X Analyzer following the manufacturer’s instructions. All plasma samples were analyzed in one batch and in duplicate with the average of the values used in subsequent analyses. Each assay included an 8-point calibrator curve, two provided kit controls, and three plasma samples from healthy individuals.

#### Validation against external normative data

We determined whether the individual plasma biomarker levels for healthy control and CAA were within or outside of age-specified normative reference intervals (RI) for plasma NfL, GFAP, p-tau-181 and the ratio of Aβ_42/40_ derived from individuals between the ages of 3 to 79 years in the Canadian Health Measures Survey (hosted by the Statistics Canada Biobank) by Cooper et al.^29^ According to each participant’s age cut-offs were defined as an upper limit (upper 95% CI of the 95^th^ percentile) for p-tau-181, NfL and GFAP while the lower limit (lower 95% CI of the 5^th^ percentile) for the Aβ_42_/Aβ_40_ ratio.^29^ These RI were generated using a different lot of the same Simoa N4PE assay and version 2.0 of the p-tau-181 assay. Thus, a conversion factor, published by Quanterix, was used to adjust the p-tau-181 values between version 2.0 and 2.1 of the assay. For participants who were 80 years of age and older, we used the published normative cut-offs for 79 years of age.^29^ Odds ratios and 95% CIs evaluate whether those with CAA had a higher, lower or equal odds of biomarker values falling outside of these normative cut offs compared to healthy controls. Additional details are provided in the **Data Supplement**.

### Statistical Analyses

Statistical analyses were conducted in Stata v18.0 (StataCorp LLC, Texas) and figures with plasma biomarker values generated using GraphPad Prism (v10.2.0). Continuous variables were evaluated for normality. Continuous variables were summarized as the mean and standard deviation (SD) if normally distributed or median and interquartile range (IQR) if not. Normally distributed variables were compared using the Student’s t-test, while non-normally distributed continuous variables were compared with the Mann-Whitney U test. Proportions of binary data were compared using the Fisher’s Exact test. Z-scores of plasma biomarker concentrations were also computed using the overall cohort mean and SD.

Differences in plasma biomarker concentrations between CAA and healthy controls, adjusting for age and sex were evaluated using linear regression. Assumptions of linear regression were evaluated and in instances where non-normality of residuals or heteroscedasticity were encountered, log transformations were applied. Binary logistic regression models were constructed to evaluate biomarker predictivity, individually and in combination, of a diagnosis of CAA accounting for age and sex. Areas under the Receiver Operating Characteristic Curve (AUC) were computed from logistic regression models to evaluate the diagnostic discriminability. AUCs of each model were compared using the test of equality of ROC areas to ascertain whether age and sex significantly improved discriminability. If an individual biomarker was significantly associated with CAA status, then it was considered for multivariable models. The Youden’s Index was computed to ascertain optimal cut points, and we report their corresponding sensitivities and specificities.

## RESULTS

### Cohort Characteristics

Overall, 92 of 120 participants had available plasma for analysis (45 CAA and 47 healthy controls). The reasons for missing samples were that samples were collected but exhausted as part of previous studies, or that there was human error in sample collection or processing. **Table 1** summarizes differences in demographic, vascular risk factor and neuroimaging variables. The first clinical presentation of CAA included lobar ICH more than 90 days prior in 16 (35.6%), TFNEs in 17 (37.8%), cognitive impairment in 10 (22.2%) and CAA-related inflammation in 2 (4.4%; these participants were studied while in remission and not taking immunosuppressants).

**Table 1:**
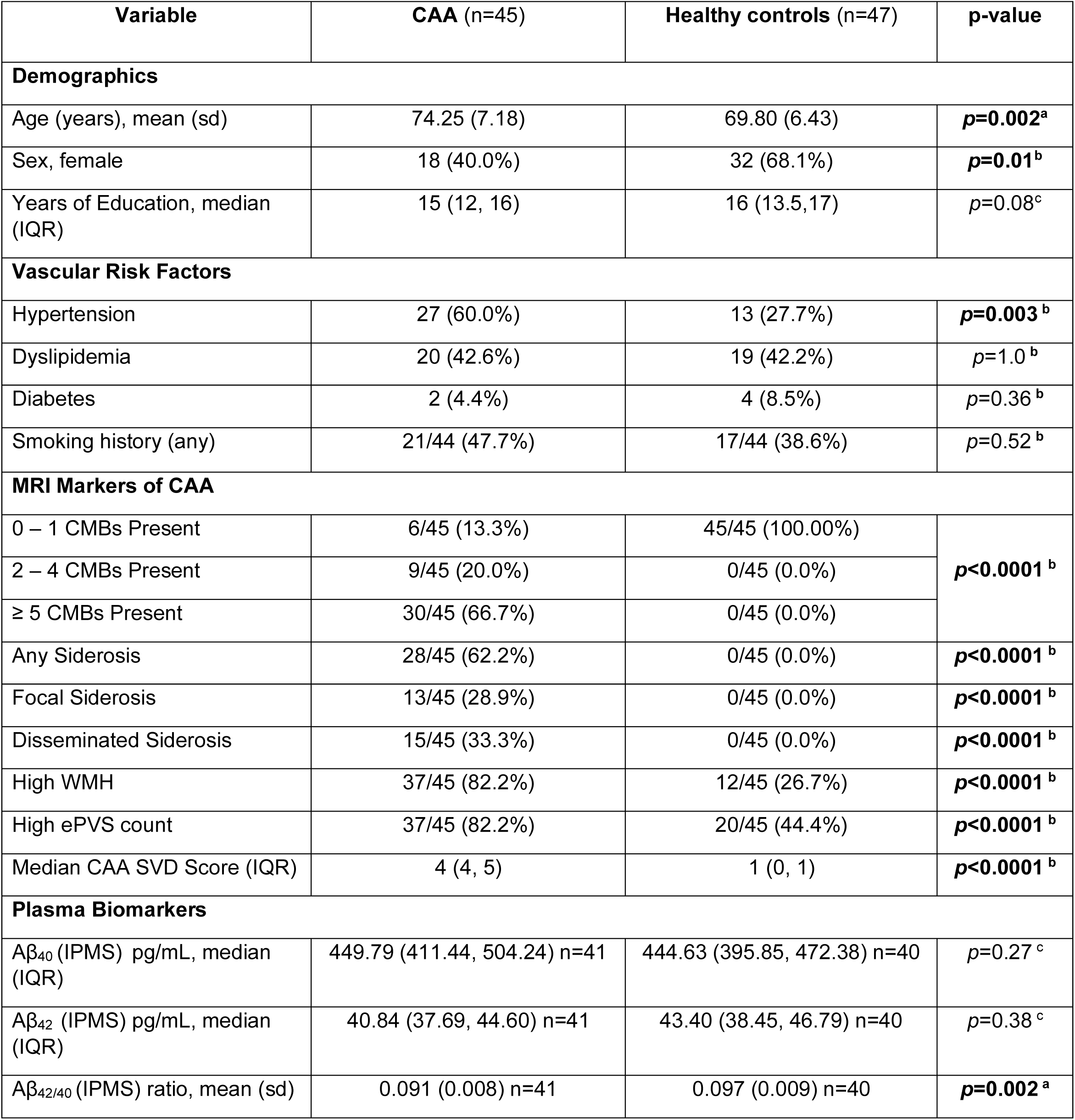
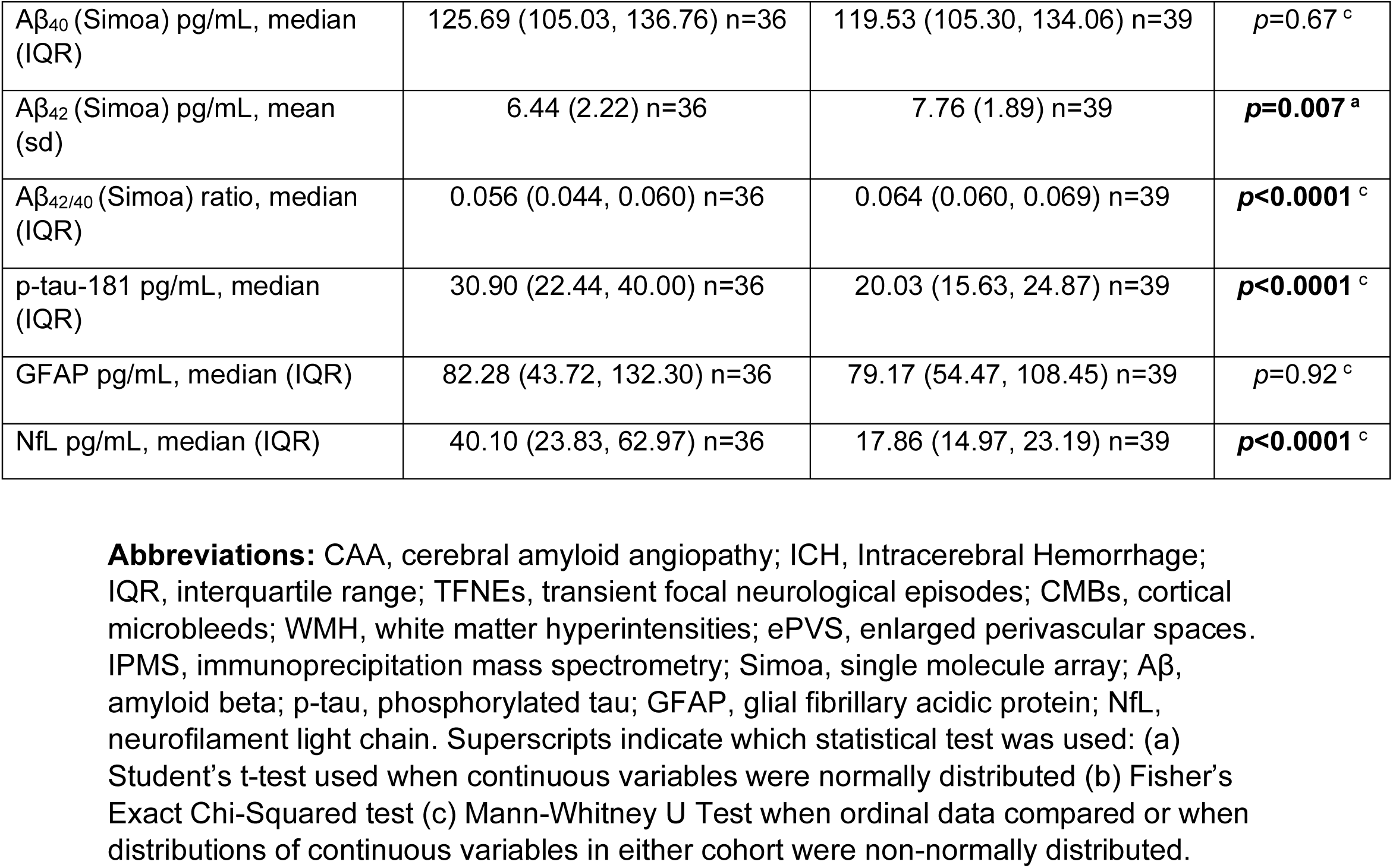
Demographic, clinical, neuroimaging and plasma biomarker characteristics of participants with CAA and healthy controls.

| Variable | CAA (n=45) | Healthy controls (n=47) | p-value |
| --- | --- | --- | --- |
| **Demographics** |  |  |  |
| Age (years), mean (sd) | 74.25 (7.18) | 69.80 (6.43) | $p=0.002^a$ |
| Sex, female | 18 (40.0%) | 32 (68.1%) | $p=0.01^b$ |
| Years of Education, median (IQR) | 15 (12, 16) | 16 (13.5,17) | $p=0.08^c$ |
| **Vascular Risk Factors** |  |  |  |
| Hypertension | 27 (60.0%) | 13 (27.7%) | $p=0.003^b$ |
| Dyslipidemia | 20 (42.6%) | 19 (42.2%) | $p=1.0^b$ |
| Diabetes | 2 (4.4%) | 4 (8.5%) | $p=0.36^b$ |
| Smoking history (any) | 21/44 (47.7%) | 17/44 (38.6%) | $p=0.52^b$ |
| **MRI Markers of CAA** |  |  |  |
| 0 – 1 CMBs Present | 6/45 (13.3%) | 45/45 (100.00%) | $p<0.0001^b$ |
| 2 – 4 CMBs Present | 9/45 (20.0%) | 0/45 (0.0%) |  |
| ≥ 5 CMBs Present | 30/45 (66.7%) | 0/45 (0.0%) |  |
| Any Siderosis | 28/45 (62.2%) | 0/45 (0.0%) | $p<0.0001^b$ |
| Focal Siderosis | 13/45 (28.9%) | 0/45 (0.0%) | $p<0.0001^b$ |
| Disseminated Siderosis | 15/45 (33.3%) | 0/45 (0.0%) | $p<0.0001^b$ |
| High WMH | 37/45 (82.2%) | 12/45 (26.7%) | $p<0.0001^b$ |
| High ePVS count | 37/45 (82.2%) | 20/45 (44.4%) | $p<0.0001^b$ |
| Median CAA SVD Score (IQR) | 4 (4, 5) | 1 (0, 1) | $p<0.0001^b$ |
| **Plasma Biomarkers** |  |  |  |
| A$\beta_{40}$ (IPMS) pg/mL, median (IQR) | 449.79 (411.44, 504.24) n=41 | 444.63 (395.85, 472.38) n=40 | $p=0.27^c$ |
| A$\beta_{42}$ (IPMS) pg/mL, median (IQR) | 40.84 (37.69, 44.60) n=41 | 43.40 (38.45, 46.79) n=40 | $p=0.38^c$ |
| A$\beta_{42/40}$ (IPMS) ratio, mean (sd) | 0.091 (0.008) n=41 | 0.097 (0.009) n=40 | $p=0.002^a$ |
| Aβ<sub>40</sub> (Simoa) pg/mL, median (IQR) | 125.69 (105.03, 136.76) n=36 | 119.53 (105.30, 134.06) n=39 | *p*=0.67<sup>c</sup> |
| Aβ<sub>42</sub> (Simoa) pg/mL, mean (sd) | 6.44 (2.22) n=36 | 7.76 (1.89) n=39 | ***p*=0.007<sup>a</sup>** |
| Aβ<sub>42/40</sub> (Simoa) ratio, median (IQR) | 0.056 (0.044, 0.060) n=36 | 0.064 (0.060, 0.069) n=39 | ***p*<0.0001<sup>c</sup>** |
| p-tau-181 pg/mL, median (IQR) | 30.90 (22.44, 40.00) n=36 | 20.03 (15.63, 24.87) n=39 | ***p*<0.0001<sup>c</sup>** |
| GFAP pg/mL, median (IQR) | 82.28 (43.72, 132.30) n=36 | 79.17 (54.47, 108.45) n=39 | *p*=0.92<sup>c</sup> |
| NfL pg/mL, median (IQR) | 40.10 (23.83, 62.97) n=36 | 17.86 (14.97, 23.19) n=39 | ***p*<0.0001<sup>c</sup>** |

**Abbreviations:** CAA, cerebral amyloid angiopathy; ICH, Intracerebral Hemorrhage; IQR, interquartile range; TFNEs, transient focal neurological episodes; CMBs, cortical microbleeds; WMH, white matter hyperintensities; ePVS, enlarged perivascular spaces. IPMS, immunoprecipitation mass spectrometry; Simoa, single molecule array; Aβ , amyloid beta; p-tau, phosphorylated tau; GFAP, glial fibrillary acidic protein; NfL, neurofilament light chain. Superscripts indicate which statistical test was used: (a) Student's t-test used when continuous variables were normally distributed (b) Fisher's Exact Chi-Squared test (c) Mann-Whitney U Test when ordinal data compared or when distributions of continuous variables in either cohort were non-normally distributed.

Of the 92 samples sent to C_2_N, 10 had insufficient volume for analysis and one had interfering substances which prevented quantification. Specific characteristics of the participants with and without available plasma, those analyzed by IPMS (n=81), Simoa (n=75) and both (n=66) are displayed in **Supplementary Tables 1-5** and patient flow diagrams are displayed in **Supplementary Figures 1 and 2**.

### Plasma Biomarker Differences

The mean (SD) or median (IQR) concentrations (pg/mL) of Aβ_40_, Aβ_42_ and the Aβ_42/40_ ratio, p-tau-181, GFAP, and NfL in CAA and healthy control groups are shown in **Table 1**.

#### Amyloid Beta isoforms

The Aβ_42/40_ ratio was lower in CAA than healthy controls with both IPMS and Simoa. Simoa plasma Aβ_42_ was significantly lower in CAA than healthy controls. There were no differences in plasma Aβ_40_ using IPMS or Simoa. Concentrations of Aβ_40,_ Aβ_42,_ and the Aβ_42/40_ ratio are displayed in **Table 1**. **Figure 1** depicts the dot plots of plasma Aβ concentrations. Overall, unadjusted Aβ_42/40_ (IPMS) and Aβ_42/40_ (Simoa) in CAA differed from healthy controls by -0.67 SD and -0.77 SD, respectively. (**Table 1** & **Supplementary Table 6**).

**Figure 1:**
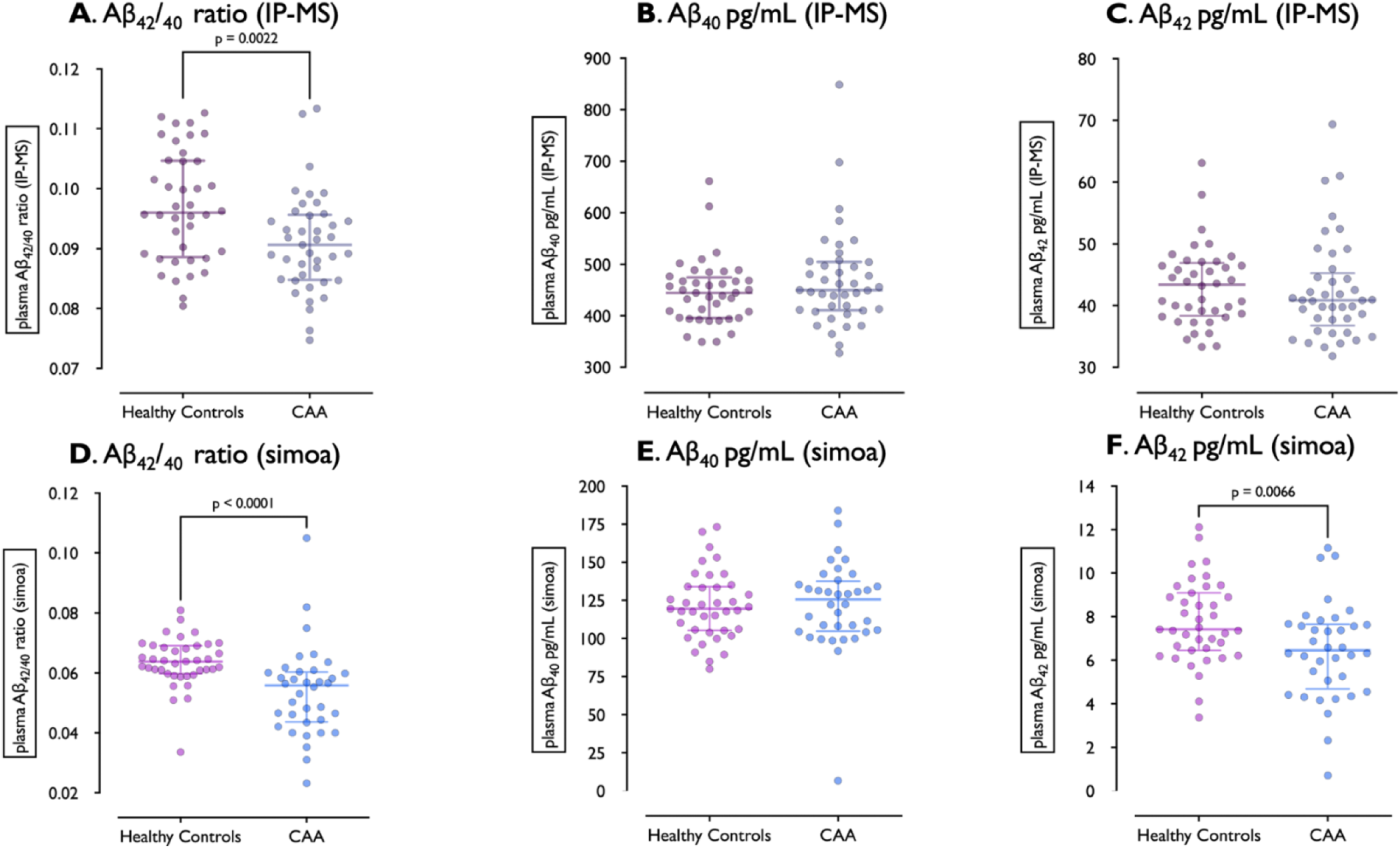
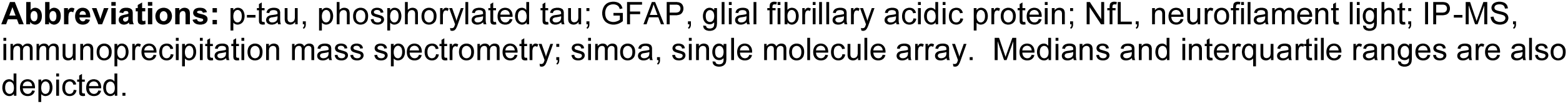
Dot plots of plasma concentrations of Aβ_40_, Aβ_42_ and the Aβ_42/40_ ratio quantified through IPMS (A-C) and Simoa (D-F) in healthy controls and CAA.

#### p-tau-181, NfL and GFAP

Participants with CAA had elevated median p-tau-181 and NfL compared to healthy controls. There was no statistically significant difference in plasma GFAP concentrations between CAA and healthy controls (**Table 1**, **Figure 2**). Overall, log(NfL) and log(p-tau-181) concentrations in CAA differed from healthy controls by +1.06 SD and +1.02 SD, respectively. (**Supplementary Table 6**).

**Figure 2:**
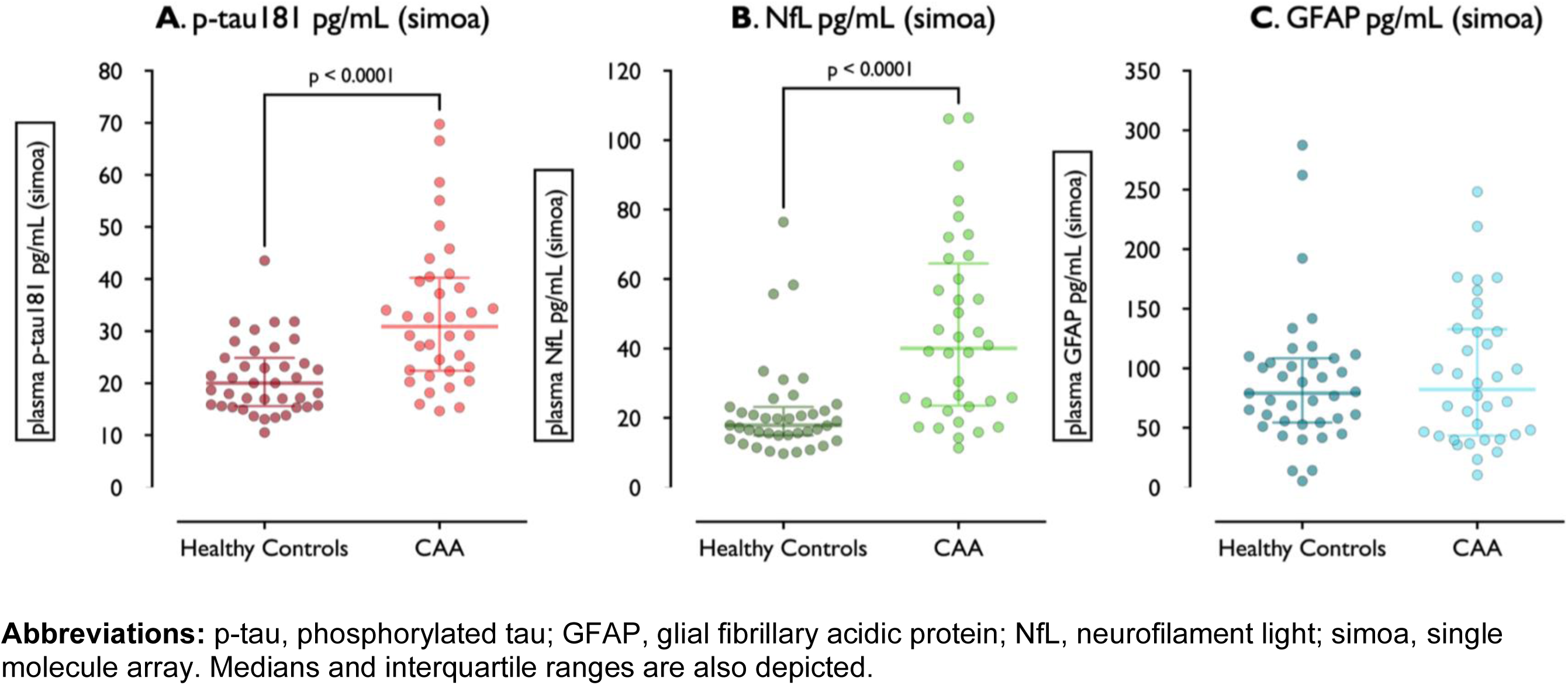
Dot plots of plasma concentrations of (A) p-tau-181 (B) NfL and (C) GFAP quantified through Simoa in healthy controls and CAA.

### Plasma biomarker differences adjusting for age and sex

Standardized beta-coefficients of the z-scores of Aβ_42/40_, log(GFAP), log(NfL), and log(p-tau-181), adjusted for age and sex, were generated in multilinear regression models. After adjustments for age and sex, significant differences in values of Aβ_42/40_ (Simoa) (β=-0.69, 95% CI: -1.15, -0.23, *p*=0.004), NfL (β=0.93, 95% CI: 0.54, 1.31, *p*<0.0001), and p-tau-181 (β=0.75, 95% CI: 0.35, 1.14, *p*<0.0001) remained, but not for Aβ_42/40_ (IPMS) (β=-0.39, 95% CI: -0.83, 0.04, *p*=0.08). (**Supplementary Table 6).**

### Validation against external normative reference intervals

Using age specific reference intervals from the Canada Health Measures Survey (n=900),^29^ participants with CAA, compared to healthy controls, had Aβ_42/40_ (Simoa), p-tau-181 (trend), NfL and GFAP levels that were more likely to fall outside of the population reference intervals (**Supplementary Table 7).**^29^ Participants with CAA had a 9.17 (95% CI: 1.07, 78.77, *p*=0.02) times greater odds than healthy controls of Aβ_42/40_ values occurring under the 5^th^ percentile as well as 10.74 (95% CI: 2.79, 41.31, *p*=0.0001) times greater odds of NfL values and 7.60 (95% CI: 0.87, 66.59, *p*=0.036) times greater odds of p-tau-181 values falling above the 95^th^ percentile compared to healthy controls (**Supplementary Table 8**). There were no differences in the proportion of those with CAA versus healthy controls with GFAP values greater than the 95^th^ percentile, however, those with CAA had a 3.84 (95% CI: 1.21, 12.25, *p*=0.02) times greater odds of GFAP values occurring less than the 5^th^ percentile compared to healthy controls (**Supplementary Table 9)**.

### Predicting a Diagnosis of Cerebral Amyloid Angiopathy (Boston Criteria 2.0)

#### Univariate Logistic Regression Analyses

**Table 2** displays the results of logistic regression models for individual biomarkers and **Figure 3A** depicts the individual ROCs for each Simoa biomarker. The **Table 3** displays the sensitivities and specificities at the Youden’s Index derived cut points for each plasma biomarker alone and in the models with subsequent age and sex adjustments.

**Figure 3:**
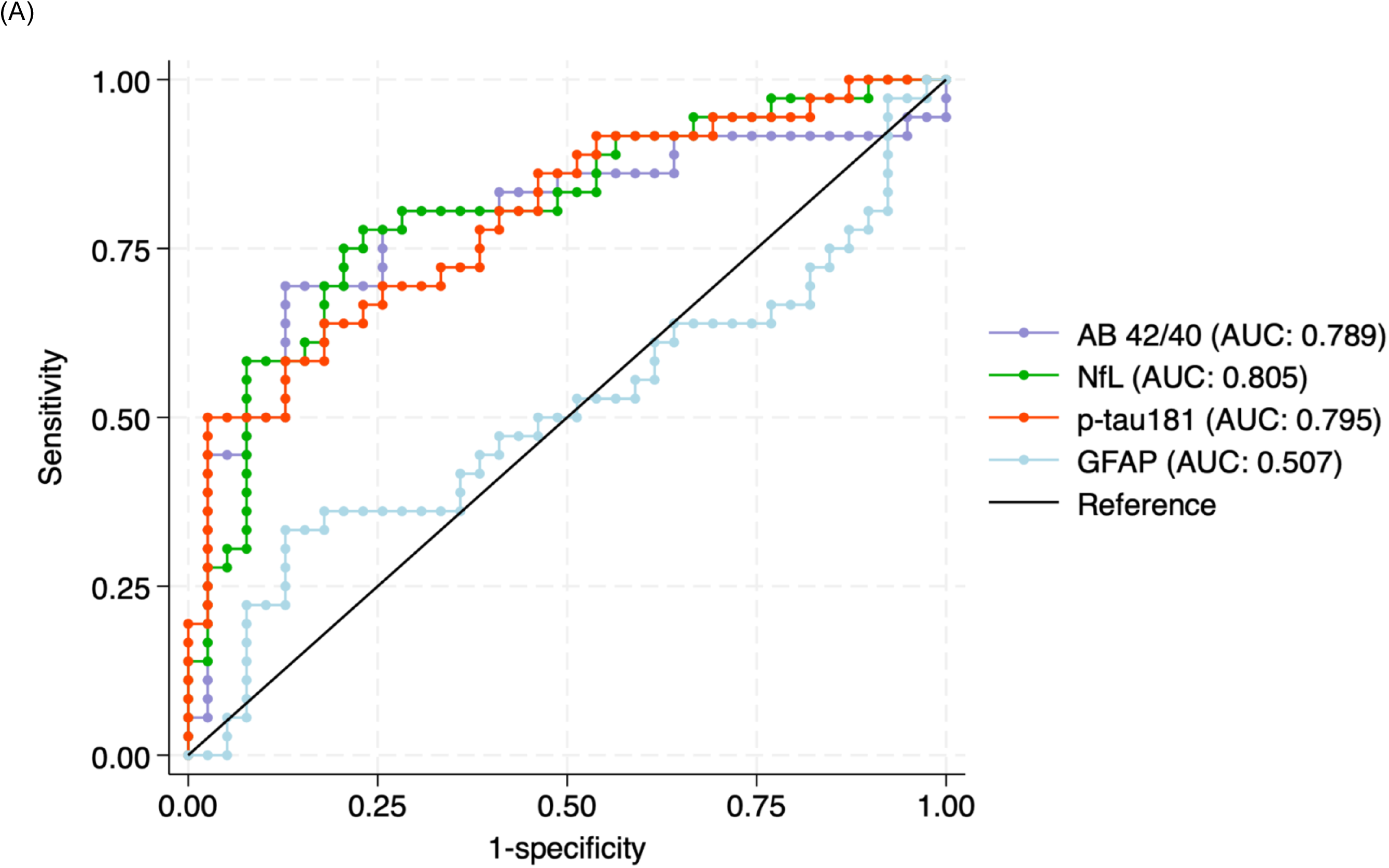
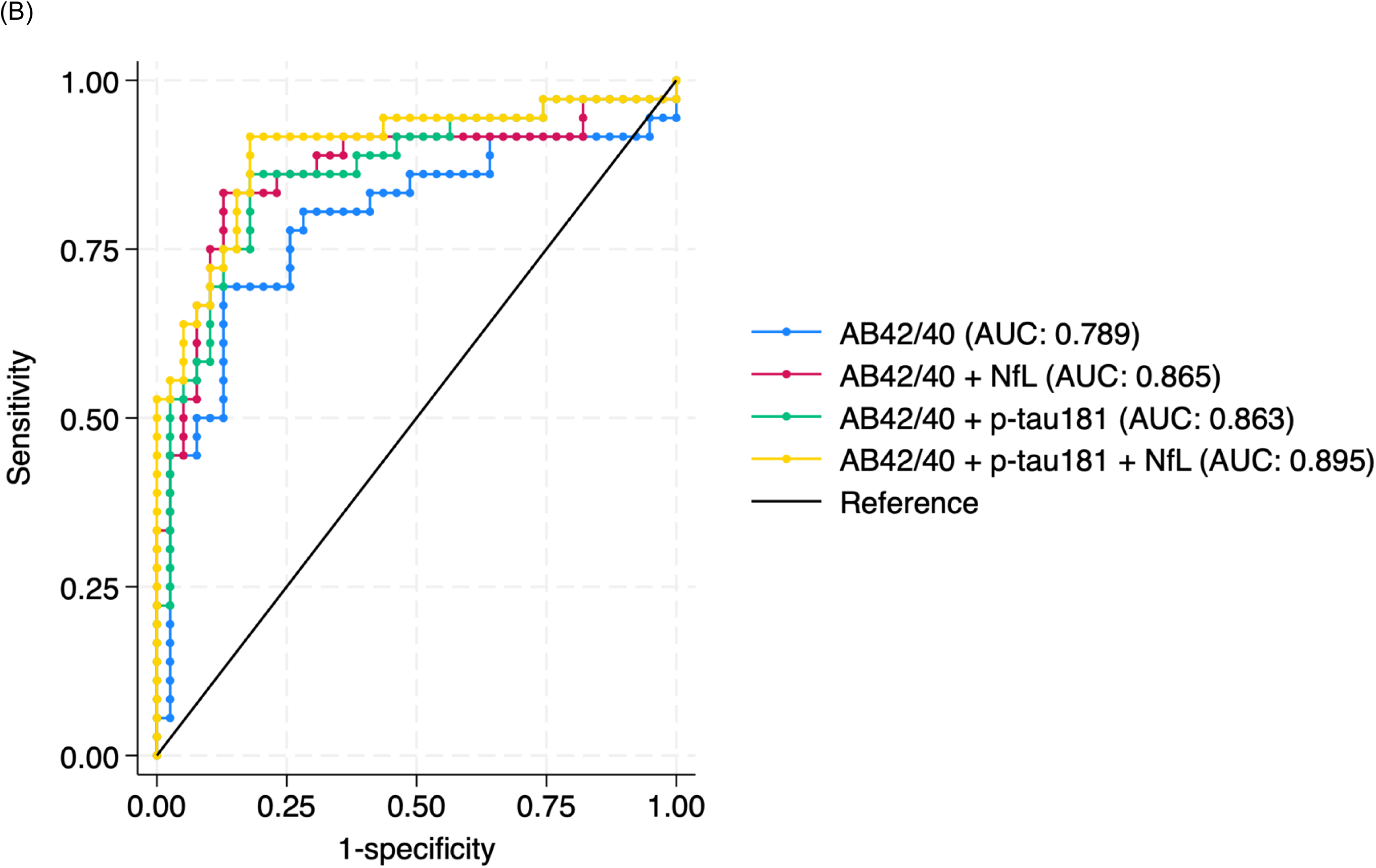
AUCs for Simoa plasma (A) Aβ_42/40_, p-tau-181, NfL and GFAP individually and (B) in combination.

**Table 2:**
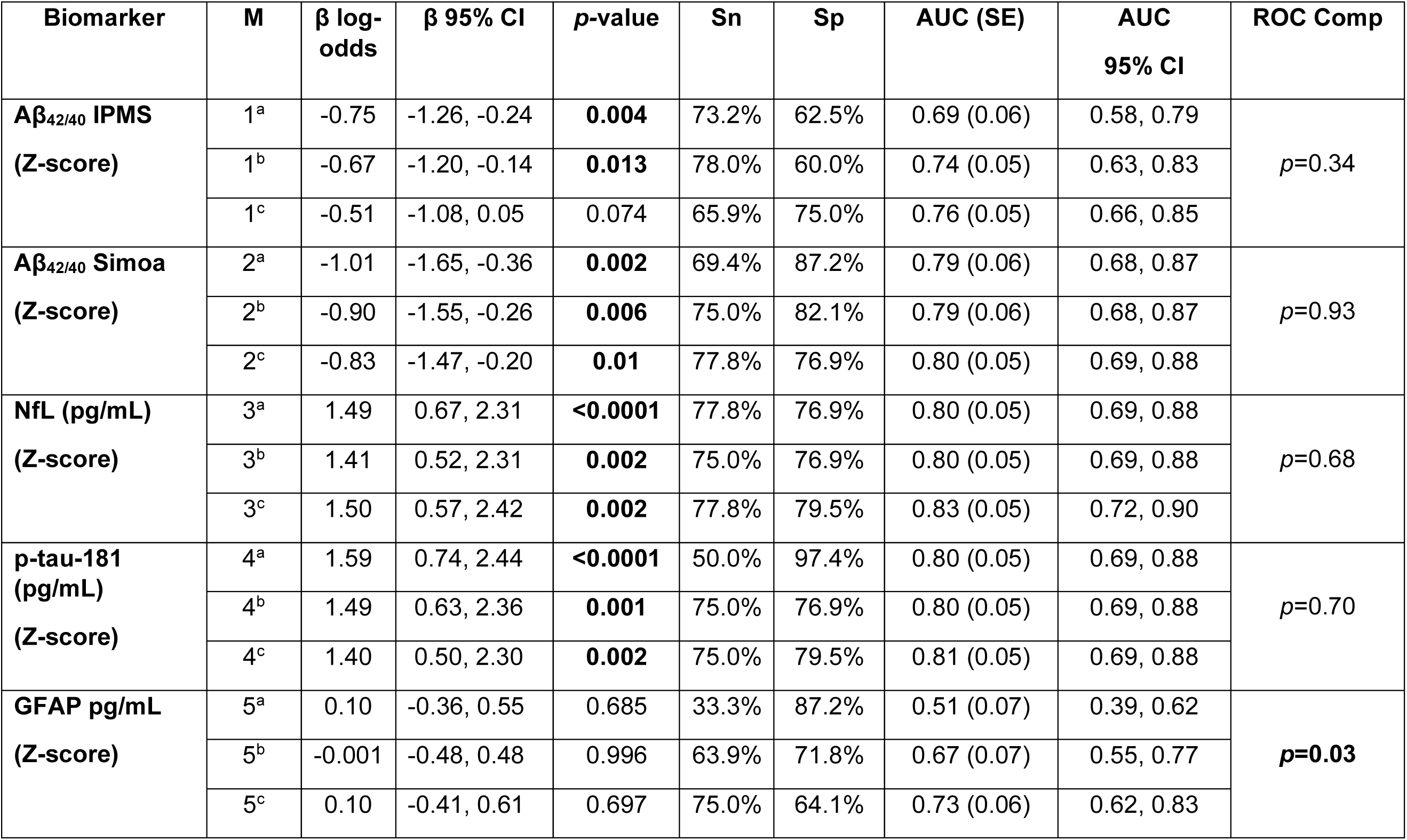
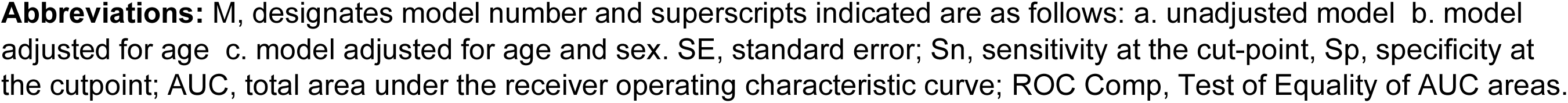
Summary of logistic regression models for individual biomarkers with standardized beta coefficients and area under the receiver operating characteristic curve (AUC)

| Biomarker | M | $\beta$ log-odds | $\beta$ 95% CI | p-value | Sn | Sp | AUC (SE) | AUC 95% CI | ROC Comp |
| --- | --- | --- | --- | --- | --- | --- | --- | --- | --- |
| **Aβ<sub>42/40</sub> IPMS**<br>**(Z-score)** | 1<sup>a</sup> | -0.75 | -1.26, -0.24 | **0.004** | 73.2% | 62.5% | 0.69 (0.06) | 0.58, 0.79 | *p*=0.34 |
|  | 1<sup>b</sup> | -0.67 | -1.20, -0.14 | **0.013** | 78.0% | 60.0% | 0.74 (0.05) | 0.63, 0.83 |  |
|  | 1<sup>c</sup> | -0.51 | -1.08, 0.05 | 0.074 | 65.9% | 75.0% | 0.76 (0.05) | 0.66, 0.85 |  |
| **Aβ<sub>42/40</sub> Simoa**<br>**(Z-score)** | 2<sup>a</sup> | -1.01 | -1.65, -0.36 | **0.002** | 69.4% | 87.2% | 0.79 (0.06) | 0.68, 0.87 | *p*=0.93 |
|  | 2<sup>b</sup> | -0.90 | -1.55, -0.26 | **0.006** | 75.0% | 82.1% | 0.79 (0.06) | 0.68, 0.87 |  |
|  | 2<sup>c</sup> | -0.83 | -1.47, -0.20 | **0.01** | 77.8% | 76.9% | 0.80 (0.05) | 0.69, 0.88 |  |
| **NfL (pg/mL)**<br>**(Z-score)** | 3<sup>a</sup> | 1.49 | 0.67, 2.31 | **<0.0001** | 77.8% | 76.9% | 0.80 (0.05) | 0.69, 0.88 | *p*=0.68 |
|  | 3<sup>b</sup> | 1.41 | 0.52, 2.31 | **0.002** | 75.0% | 76.9% | 0.80 (0.05) | 0.69, 0.88 |  |
|  | 3<sup>c</sup> | 1.50 | 0.57, 2.42 | **0.002** | 77.8% | 79.5% | 0.83 (0.05) | 0.72, 0.90 |  |
| **p-tau-181 (pg/mL)**<br>**(Z-score)** | 4<sup>a</sup> | 1.59 | 0.74, 2.44 | **<0.0001** | 50.0% | 97.4% | 0.80 (0.05) | 0.69, 0.88 | *p*=0.70 |
|  | 4<sup>b</sup> | 1.49 | 0.63, 2.36 | **0.001** | 75.0% | 76.9% | 0.80 (0.05) | 0.69, 0.88 |  |
|  | 4<sup>c</sup> | 1.40 | 0.50, 2.30 | **0.002** | 75.0% | 79.5% | 0.81 (0.05) | 0.69, 0.88 |  |
| **GFAP pg/mL**<br>**(Z-score)** | 5<sup>a</sup> | 0.10 | -0.36, 0.55 | 0.685 | 33.3% | 87.2% | 0.51 (0.07) | 0.39, 0.62 | ***p*=0.03** |
|  | 5<sup>b</sup> | -0.001 | -0.48, 0.48 | 0.996 | 63.9% | 71.8% | 0.67 (0.07) | 0.55, 0.77 |  |
|  | 5<sup>c</sup> | 0.10 | -0.41, 0.61 | 0.697 | 75.0% | 64.1% | 0.73 (0.06) | 0.62, 0.83 |  |

**Abbreviations:** M, designates model number and superscripts indicated are as follows: a. unadjusted model b. model adjusted for age c. model adjusted for age and sex. SE, standard error; Sn, sensitivity at the cut-point, Sp, specificity at the cutpoint; AUC, total area under the receiver operating characteristic curve; ROC Comp, Test of Equality of AUC areas.

**Table 3:** Summary of logistic regression model receiver operating characteristic curves (AUC) for combinations of plasma biomarkers.

| Aβ<sub>42/40</sub> IPMS + Simoa biomarkers (n=66) | Sn | Sp | AUC (SE) | AUC 95% CI | ROC Comp |
| --- | --- | --- | --- | --- | --- |
| Aβ<sub>42/40</sub> | 73.2% | 62.5% | 0.66 (0.07) | 0.52, 0.77 | p=0.01 |
| Aβ<sub>42/40</sub> + NfL | 75.8% | 87.9% | 0.86 (0.05) | 0.76, 0.94 |  |
| Aβ<sub>42/40</sub> + p-tau-181 | 48.5% | 97.0% | 0.77 (0.06) | 0.65, 0.87 |  |
| Aβ<sub>42/40</sub> + NfL + p-tau-181 | 78.8% | 87.9% | 0.87 (0.05) | 0.76, 0.94 | p=0.72 |
| Aβ<sub>42/40</sub> + NfL + p-tau-181 + age + sex | 78.8% | 87.9% | 0.88 (0.04) | 0.78, 0.95 |  |
|  | 75.8% | 90.9% |  |  |  |
| Aβ<sub>42/40</sub> Simoa + Simoa biomarkers (n=75) | Sn | Sp | AUC (SE) | AUC 95% CI | ROC Comp |
| Aβ<sub>42/40</sub> | 69.4% | 87.2% | 0.79 (0.06) | 0.68, 0.87 | p=0.08 |
| Aβ<sub>42/40</sub> + NfL | 83.3% | 87.2% | 0.87 (0.05) | 0.77, 0.93 |  |
| Aβ<sub>42/40</sub> + p-tau-181 | 86.1% | 82.1% | 0.86 (0.05) | 0.77, 0.93 |  |
| Aβ<sub>42/40</sub> + NfL + p-tau-181 | 91.7% | 82.1% | 0.90 (0.04) | 0.80, 0.95 | p=0.71 |
| Aβ<sub>42/40</sub> + NfL + p-tau-181 + age + sex | 86.1% | 79.5% | 0.90 (0.04) | 0.82, 0.96 |  |
| Simoa biomarkers without Aβ<sub>42/40</sub> (n=75) | Sn | Sp | AUC (SE) | AUC 95% CI | ROC Comp |
| NfL + p-tau-181 | 88.9% | 66.7% | 0.84 (0.04) | 0.74, 0.91 | p=0.12* |

**Abbreviations:** Sn, sensitivity at the Youden's Index defined cut-point, Sp, specificity at the Youden's Index defined cutpoint; AUC, total area under the receiver operating characteristic curve; ROC Comp, Test of Equality of AUC areas. Note: Where ties occurred a single Youden's Index could not be computed and thus two probability cut-points are reported with maximal correctly classified participants. \* note this p-value is for the test of equality of AUC areas comparing this model to the model A $\beta_{42/40}$ + NfL + p-tau-181.

Individually, Aβ_42/40_ (IPMS), Aβ_42/40_ (Simoa), NfL, and p-tau-181 all predicted a diagnosis of CAA with AUCs of 0.69 for Aβ_42/40_ (IPMS) (95% CI: 0.58, 0.79, cut-point: 0.095), 0.79 for Aβ_42/40_ (Simoa) (95% CI: 0.68, 0.87, cut-point: 0.059); 0.81 for NfL (95% CI: 0.69, 0.88, cut-point: 23.22pg/mL), and 0.80 for p-tau-181 (95% CI: 0.69, 0.88, cut-point: 32.24 pg/mL). The addition of age and sex to these models did not increase their discriminative performance. The most specific single biomarker was p-tau-181 with a specificity of 97.4% and sensitivity of 50.0%, while the most sensitive single biomarker was NfL with a sensitivity of 77.8% and specificity of 76.9%. Aβ_42/40_ had intermediate sensitivities and specificities. GFAP alone did not discriminate a diagnosis of CAA from healthy controls.

#### Multivariable Logistic Regression Analyses

Multiple biomarkers were evaluated in multivariable logistic regression models to compute AUCs, Youden’s Index derived probability cut points and corresponding sensitivities and specificities at these cut-points (**Table 3**). The addition of NfL and p-tau-181 to a base model with Aβ_42/40_ (IPMS) improved the discriminative performance (*p*=0.01), but the addition of NfL and p-tau-181 to a base model with Aβ_42/40_ (Simoa) did not (*p*=0.08).

Overall convergent results were noted across the Aβ_42/40_ methods used. In the subset with both Aβ_42/40_ (IPMS) and Simoa based p-tau-181 and NfL, these plasma biomarkers predicted a diagnosis of CAA with an AUC of 0.87 (95% CI: 0.76, 0.94), with a sensitivity of 78.8% and specificity of 87.9% at the probability cut-point.

In the subset with Simoa measured biomarkers, combining Aβ_42/40_, p-tau-181, and NfL yielded the most sensitive combination of biomarkers and predicted a diagnosis of CAA with an AUC 0.90 (95% CI: 0.80, 0.95), with a sensitivity of 91.7% and specificity of 82.1% at the probability cut-point. The most specific combination of biomarkers, on the other hand, was observed with Aβ_42/40_ (IPMS) and p-tau-181 (sensitivity: 48.5%, specificity: 97.0%). The ROCs of the multivariable logistic regression models with Simoa based biomarkers are displayed in **Figure 3B**. The addition of age and sex to these models did not increase their discriminative performance (*p*≥0.7). **Supplementary Table 10** depicts the logistic regression model log-odds beta-coefficients along with cut-points derived from Youden’s Index.

## DISCUSSION

Participants with Boston Criteria 2.0 defined probable CAA had reduced plasma Aβ_42/40_ ratios and elevated p-tau-181 and NfL concentrations compared to healthy controls. A combination of Aβ_42/40_ (Simoa), p-tau-181, and NfL resulted in the highest AUC of 0.90 (95% CI: 0.80, 0.95) and a corresponding sensitivity and specificity of 91.7% and 82.1%. The diagnostic test performance of these three plasma biomarkers is considered excellent.

These data raise some interesting pathophysiologic questions about CAA. The differences in plasma biomarkers we observed between CAA and healthy controls is a pattern of change that may also occur with AD, except that studies have reported elevations in GFAP in AD,^30, 31^ while we found that GFAP was not different in CAA. While our participants do have probable CAA, we cannot exclude the possibility of some degree of concomitant AD pathology. Asymptomatic cerebral amyloidosis is common with increasing age,^15^ and since we did not screen our controls for amyloidosis before enrollment it is possible that intermediate sensitivities and specificities noted with Aβ_42/40_ could be related to asymptomatic amyloidosis, which could bias our results towards null differences.

Elevations in p-tau-181 in our study may be because of concomitant AD with CAA, or may be due to direct effects of CAA on tau levels. Plasma p-tau is not just a marker of cerebral tau status, but is also an indicator of cerebral amyloid deposition.^32^ Neuropathological evidence suggests that tau accumulation occurs in the neurites proximal to vessels affected by CAA.^33, 34^ Our plasma results are concordant with those from CSF, where patients with CAA have elevated CSF t-tau and p-tau compared to controls, but not as high as those with AD.^35^ In an another neuropathological study the correlation between tau-tangles and amyloid was higher for amyloid plaque pathology (r=0.77) than amyloid angiopathy (r=0.32).^36^ Another analysis of 1722 individuals found that those with greater severities of CAA and plaque pathology had higher tau pathology, suggesting that tau burden may jointly depend on both CAA and plaque pathology.^37^

Increased GFAP in AD may be due to reactive astrocytosis to parenchymal amyloid plaque and, furthermore, GFAP can predict the conversion from amnestic MCI to AD.^38, 39^ In a neuropathological study, plasma GFAP was elevated in those with greater neuritic plaque pathology but not with CAA.^40^ Therefore, the comparable plasma concentrations of GFAP between CAA and healthy controls observed in our cohort might be a result of a lower burden of amyloid plaque in our CAA cohort. Additional interpretations of this finding might be that: (i) astrocytosis may not be as prominent feature of CAA; (ii) there is impaired clearance of GFAP into systemic circulation; or (iii) there is retreat of glial end feet from vessels afflicted with CAA. Larger studies are needed in the future to confirm whether a combination of p-tau and GFAP can discriminate CAA (high p-tau, normal GFAP) from AD (higher p-tau, high GFAP).

There are some important differences and similarities with respect to our findings in plasma and studies of CSF in CAA. A meta-analysis of CSF biomarkers reports elevations in t-tau, reductions in Aβ_42_ and Aβ_40_, and marginal differences with p-tau in CAA compared to controls.^41^ Compared with AD, patients with CAA had lower Aβ_40_, similar Aβ_42_, lower p-tau and lower t-tau.^41^ One cohort study of CAA (n=67), AD (n=76), mild cognitive impairment (MCI) related to AD (n=75), MCI unrelated to AD (n=76), and controls (n=78) noted the following patterns of CSF biomarker profiles: (i) reductions in CSF Aβ_40_ in CAA compared to controls, AD and MCI-AD, and (ii) elevations in t-tau and p-tau in CAA compared to MCI-non-AD and controls, but lower than AD and MCI-AD.^42^ In plasma we did not observe differences between CAA and healthy controls in Aβ_40_ using either Simoa or IPMS methodology. Ultimately, CSF Aβ_40_ may be a more accurate reflection of central nervous system CAA than plasma Aβ_40_. However, we did find find that adding Aβ_42/40_ to a model with p-tau-181 and NfL increased the specificity from 67% to 82%, suggesting that measuring plasma Aβ_42/40_ does have utility. One review did not find differences in either plasma Aβ_40_ or Aβ_42_ in those with CAA compared to controls, but there was heterogeneity across plasma quantification methods and the Aβ_42/40_ ratio was not examined.^43^ In a large cohort study of participants with CAA related-ICH, 95 healthy controls were compared to participants with probable CAA related acute ICH (n=68) and, similar to our study, using Simoa methodology, elevations in plasma NfL and a reduction in the plasma Aβ_42/40_ ratio were noted.^44^ Interestingly, plasma NfL predicted future ICH recurrence.^44^ There have been a small number of other studies to date that evaluated plasma biomarkers in sporadic and autosomal dominant CAA. In a recent study there were no differences in plasma Aβ_40_ or Aβ_42_ in 61 cases of sporadic CAA compared to 42 matched controls, but in those with symptomatic hereditary Dutch type CAA, plasma Aβ_42_ levels were significantly lower compared to controls.^45^ This study did not examine the Aβ_42/40_ ratio. Another smaller study in mutation carriers for Dutch-type CAA noted reductions in plasma Aβ_40_, and Aβ_42_ across time points.^46^ Ultimately, there has been variability in results across small studies in participants with CAA examining plasma Aβ_40_ and/or Aβ_42_ with different quantification methods.^47-49^

A strength of our study is that we corroborated the observations of the Aβ_42/40_ ratio using two validated modern methodologies: Simoa and IPMS. Furthermore, we leverage the existence of reference intervals for Simoa plasma biomarkers in the Canadian population to show that those with CAA, compared to healthy controls, had a 7.6 to 10.7 times greater odds of having plasma Aβ_42/40_, p-tau-181, and NfL values falling outside of reference intervals. This supports our study’s external validity.

Limitations of this study include the small sample size and lack of external validation. Furthermore, including participants with neuropathological diagnosis of CAA, is needed to confirm the findings. For initial discovery, we used the Boston Criteria 2.0 as a reference standard, which has a sensitivity of 79.8% and specificity of 84.7% compared to neuropathologically confirmed CAA.^4^ Some of the apparent false positive and false negatives in this study may be due to the imperfect accuracy of the Boston Criteria 2.0, rather than limitations of the plasma assays, or possibly due to concomitant AD. Future analyses should also consider including APOE genotype in predicting CAA.^50^ Finally, the previously published external normative reference intervals were run on a different lot and, thus, lot variability may be a source of bias.^29^

Overall, this study suggests that plasma Aβ_42/40_, p-tau-181 and NfL together can discern Boston Criteria 2.0 probable CAA from healthy controls. Compared to PET and CSF studies, plasma is more accessible and scalable. Identifying a plasma biomarker signature of CAA may advance the diagnostic discriminability beyond clinical and imaging criteria and may also facilitate the diagnostic workup for those with ICH and those who cannot undergo MRI. Future studies should compare plasma markers profiles in CAA to AD and non-CAA ICH and determine whether plasma markers can predict recurrent ICH. Our study also highlights the need for investigations of biomarkers in larger cohorts of CAA, which may be expeditiously leveraged by international collaborative working groups with pooled test and validation cohorts. Considering the recent advances in AD beta-amyloid targeting immunotherapies and the concern raised for a higher risk of ARIA among those with concomitant CAA, identifying a distinguishing plasma biomarker panel of CAA from AD will be of paramount importance in future.^12, 13, 19^

## DATA SHARING STATEMENT

Anonymized data will be made available to other qualified researchers on reasonable request to the corresponding author for the purposes of replicating analyses.

## ACKNOWLEDGEMENTS

The study was funded by Dr. Eric E Smith’s Canadian Institutes of Health Research (CIHR) operating grant. Dr. Ryan T. Muir gratefully acknowledges support from the CIHR, the Hotchkiss Brain Institute at the University of Calgary, and the Alzheimer’s Society of Canada. C_2_N Diagnostics has received support from the NIH (grant No. R44 AG059489), BrightFocus (grant No. CA2016636), The Gerald and Henrietta Rauenhorst Foundation, and the Alzheimer’s Drug Discovery Foundation (grant No. GC-201711-2013978).

## DATA SUPPLEMENT

### Supplementary Methodology

#### Plasma Biomarker Quantification

##### Simoa

Quality control measures were recorded, and all samples fell within the assay limits of quantification. There were 4 samples with high intra-sample coefficients of variability (CV) for p-tau-181 (CVs>50%) unrelated to machine or sample quality. These four samples did not pose a problem for the N4PE assay, which implied that there was not an overall problem with the sample. These were re-run and CVs<10.18% were achieved upon re-analysis. The values from the repeated analyses were valid and subsequently used.

While normative data from the ageing Canadian population as reported in Cooper et al.^29^ for plasma Aβ_42/40_, GFAP, and NfL were based off the Neurology-4-plex E advantage assay (catalogue 103670, lot 503105) version 2.1 Advantage Kit (cat ID 104111, lot 503843) was used, the plasma p-tau-181 was measured using the p-tau-181 version 2.0 advantage assay (catalogue 103714, lot 502923). In the current analyses the version 2.1 Advantage Kit (catalogue #104111, lot 503843) was used. Therefore, to compare v2.1 p-tau-181 concentrations in the current analyses to the normative data from Cooper et al.^29^ a conversion factor published by Quanterix was applied. This conversion factor is published in a technical note by Quanterix entitled: Simoa p-tau 181 Advantage V2.1 Assay (reference TECH-0153), released specifically to describe the differences between the V2.0 and V2.1 assay. ^51^

#### Statistical Analyses

##### Linear Regression

Multivariable linear regression models were constructed to evaluate for differences in plasma biomarkers between groups while adjusting for age and sex. The plasma biomarker values were transformed to Z scores by subtracting the group mean from each participant’s value and dividing this by the group standard deviation. In all models the assumptions of no multicollinearity, normality of residuals, and homoscedasticity were evaluated. Normality of residuals were evaluated with kernel density curves and q-q plots. Homoscedasticity was evaluated using Cameron & Trivedi’s test for heteroskedasticity. Multi-collinearity was evaluated using Variance Inflation Factor. In instances where the assumptions of normality of residuals and/or homoscedasticity were violated, the plasma biomarker values were log transformed first then transformed to Z scores for modelling. The results of these models are reported below in **Supplementary Table 7.**

#### Comparison to normative population data

As the data provided in Cooper et al.^29^ apply to an age range of 3 to 79 years and our dataset had participants 80 and over, we used the 79 year-old reference values for the 9 participants age 80 and over. To define those <5^th^ percentile the lower bound 95% confidence interval value of the predicted age specific 5^th^ percentile value was used. To define those >95^th^ percentile the upper bound of the 95% confidence interval value of the predicted age specific 95^th^ percentile value was used. These data are displayed below in **Supplementary Table 7**.

The number of participants in the Normal Control and CAA cohorts with (A) within the range of normative values produced from Cooper et al were “Test Negative” and (B) out of range, “Test Positive,” abnormal was defined as (1) <5%tile for Aβ_42/40_ (2) >95%tile for p-tau-181, GFAP, and NfL. An odds ratio was then constructed using the Woolf method for estimating confidence intervals around the point estimate (as displayed in **Supplementary Table 7**).

One of the important limitations for using the population normative data from Cooper et al. ^29^, is that the the recruited n=900 participants were consenting Canadians between the ages of 3 – 79 years without any inclusion or exclusion criteria on the basis of cognitive impairment or the presence of neurologic disorders which can lead to cognitive impairment and/or dementia. It is likely that the normative population data from Cooper et al. contains persons with cognitive disorders and concomitant neurologic disorders (such as neurodegenerative diseases). Thus, the normative population data are not equivalent to our healthy control cohort where exclusion criteria were in place for cognitive impairment and concomitant neurological disorders.

## Supplementary Tables

**Supplementary Table 1:**
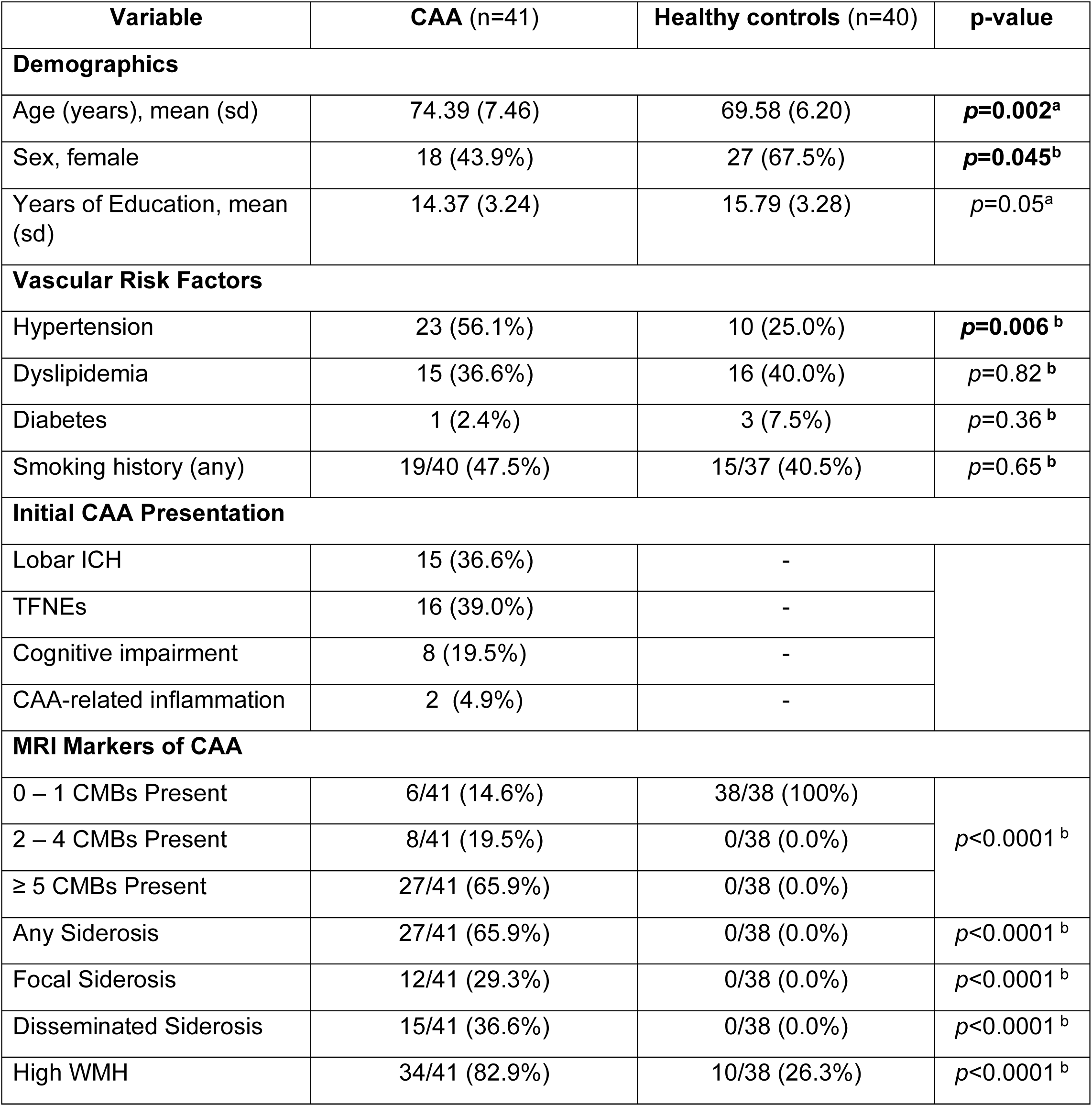
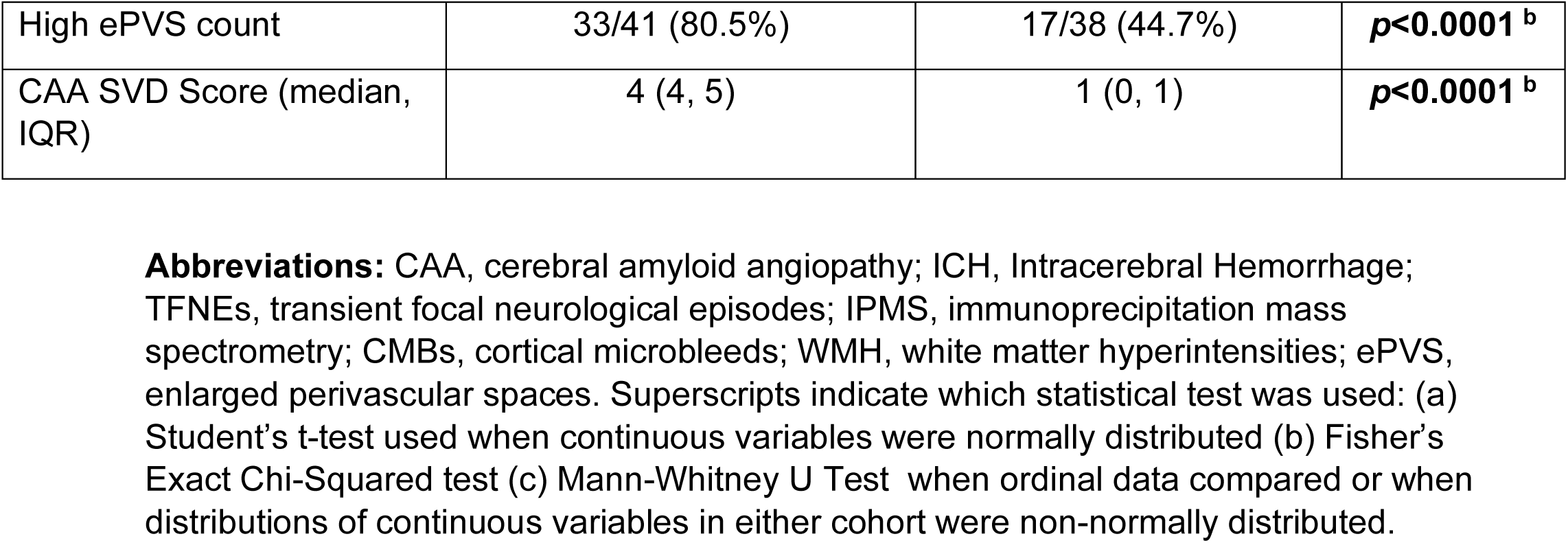
Demographic, Vascular Risk Factor, Cognitive and Neuroimaging Data for participants with sufficient samples sent to C2N for IPMS quantification of plasma amyloid-beta.

**Supplementary Table 2:** Demographic, Vascular Risk Factor, Cognitive and Neuroimaging Data for participants with sufficient samples sent for Simoa quantification plasma amyloid-beta, p-tau-181, NfL, and GFAP.

| Variable | CAA (n=36) | Healthy controls (n=39) | p-value |
| --- | --- | --- | --- |
| **Demographics** |  |  |  |
| Age (years), mean(sd) | 74.11 (7.29) | 70.39 (6.10) | ***p*=0.02<sup>a</sup>** |
| Sex, female | 16 (44.4%) | 27 (69.2%) | ***p*=0.04<sup>b</sup>** |
| Years of Education, median (IQR) | 15 (12.5, 17) | 15 (13.5, 17) | *p*=0.50<sup>c</sup> |
| **Vascular Risk Factors** |  |  |  |
| Hypertension | 21 (58.3%) | 10 (25.6%) | ***p*=0.005<sup>b</sup>** |
| Dyslipidemia | 14 (38.89%) | 16 (41.0%) | *p*=1.00<sup>b</sup> |
| Diabetes | 1 (2.8%) | 3 (7.7%) | *p*=0.62<sup>b</sup> |
| Smoking history (any) | 13/35 (37.1%) | 14/37 (37.8%) | *p*=1.00<sup>b</sup> |
| **Initial CAA Presentation** |  |  |  |
| Lobar ICH | 13 (36.1%) | - |  |
| TFNEs | 14 (38.9%) | - |  |
| Cognitive impairment | 7 (19.4%) | - |  |
| CAA-related inflammation | 2 (5.6%) | - |  |
| **MRI Markers of CAA** |  |  |  |
| 0 – 1 CMBs Present | 5 (13.9%) | 38/38 (100.0%) | ***p*<0.0001<sup>b</sup>** |
| 2 – 4 CMBs Present | 7 (19.4%) | 0/38 (0.0%) |  |
| ≥ 5 CMBs Present | 24 (66.7%) | 0/38 (2.6%) |  |
| Any Siderosis | 23 (63.9%) | 0/38 (0.0%) | ***p*<0.0001<sup>b</sup>** |
| Focal Siderosis | 11 (30.6%) | 0/38 (0.0%) | ***p*<0.0001<sup>b</sup>** |
| Disseminated Siderosis | 12 (33.3%) | 0/38 (0.0%) | ***p*<0.0001<sup>b</sup>** |
| High WMH | 29 (80.6%) | 11/38 (29.0%) | ***p*<0.0001<sup>b</sup>** |
| High ePVS count | 30 (83.3%) | 19/38 (50.0%) | ***p*=0.003<sup>b</sup>** |
| CAA SVD Score (median, IQR) | 4 (4, 5) | 1 (0, 1) | ***p*<0.0001<sup>b</sup>** |

**Abbreviations:** CAA, cerebral amyloid angiopathy; ICH, Intracerebral Hemorrhage; TFNEs, transient focal neurological episodes; Simoa, single molecule array; Aβ , amyloid beta; p-tau, phosphorylated tau; GFAP, glial fibrillary acidic protein; NfL, neurofilament light; CMBs, cortical microbleeds; WMH, white matter hyperintensities; ePVS, enlarged perivascular spaces. Superscripts indicate which statistical test was used: (a) Student's t-test used when continuous variables were normally distributed (b) Fisher's Exact Chi-Squared test (c) Mann-Whitney U Test when ordinal data compared or when distributions of continuous variables in either cohort were non-normally distributed.

**Supplementary Table 3:**
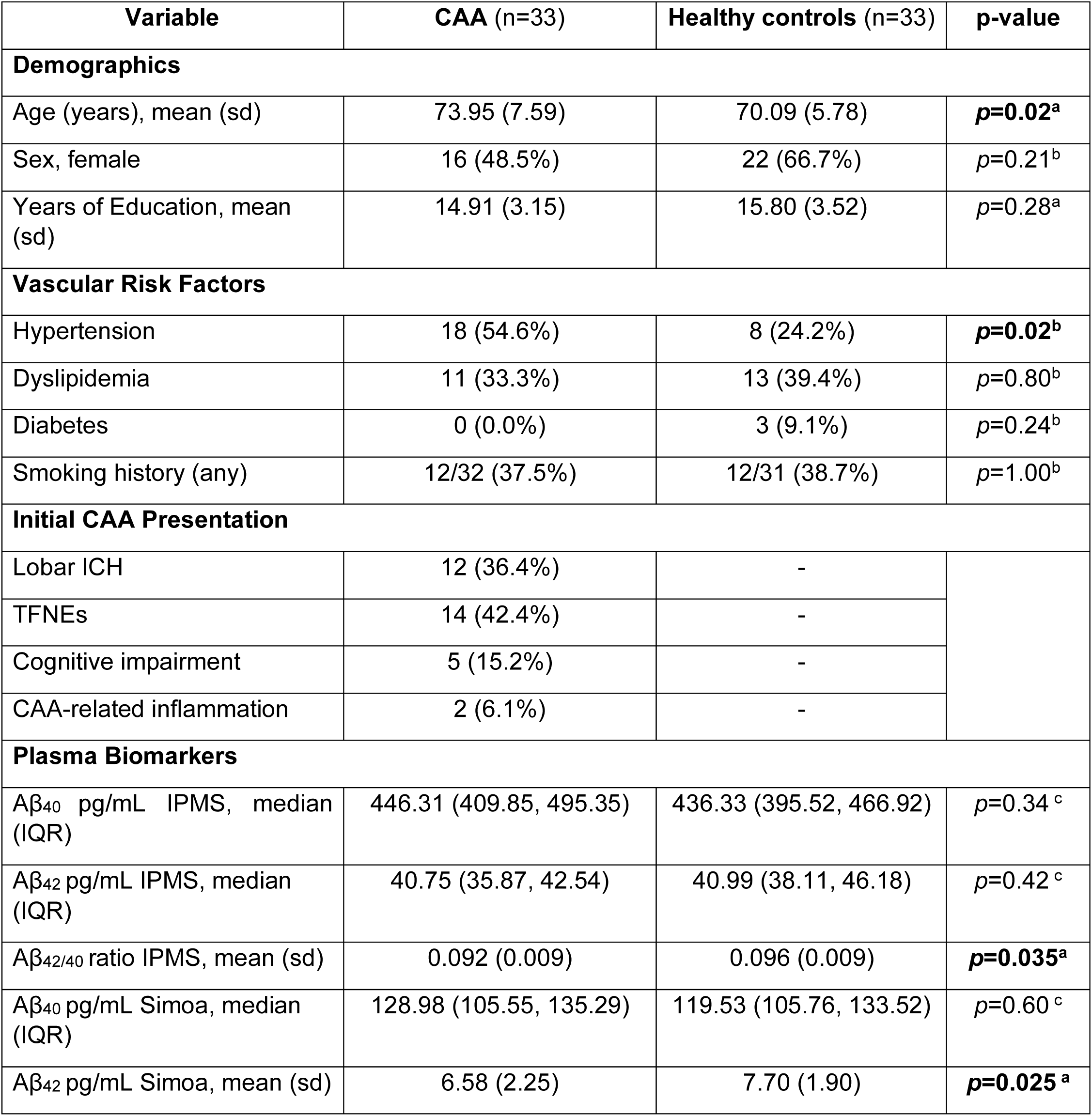
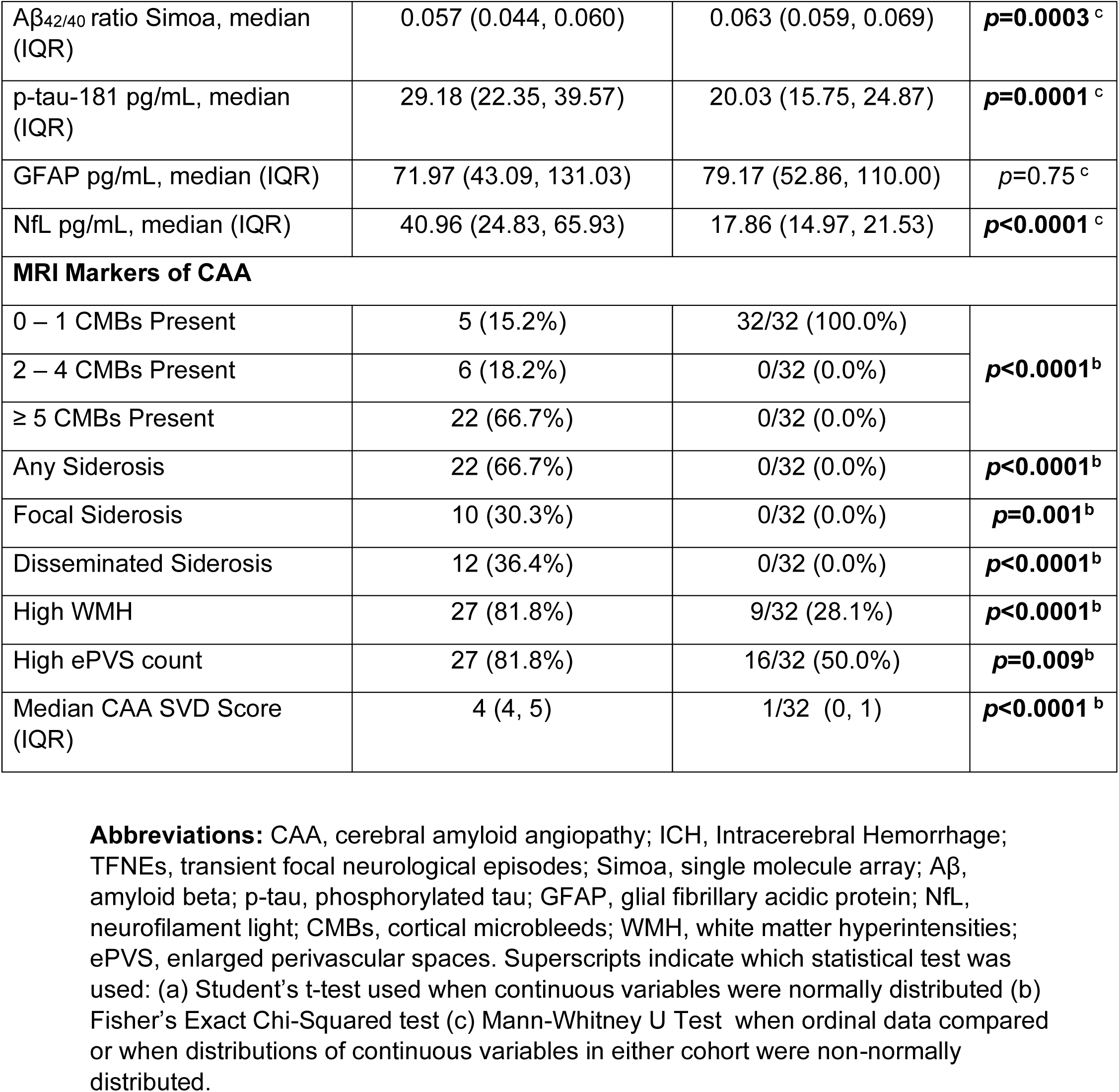
Demographic, Vascular Risk Factor, Plasma Biomarker, Cognitive and Neuroimaging Data for participants who had both IPMS and Simoa plasma biomarker quantification.

**Supplemental Table 4:**
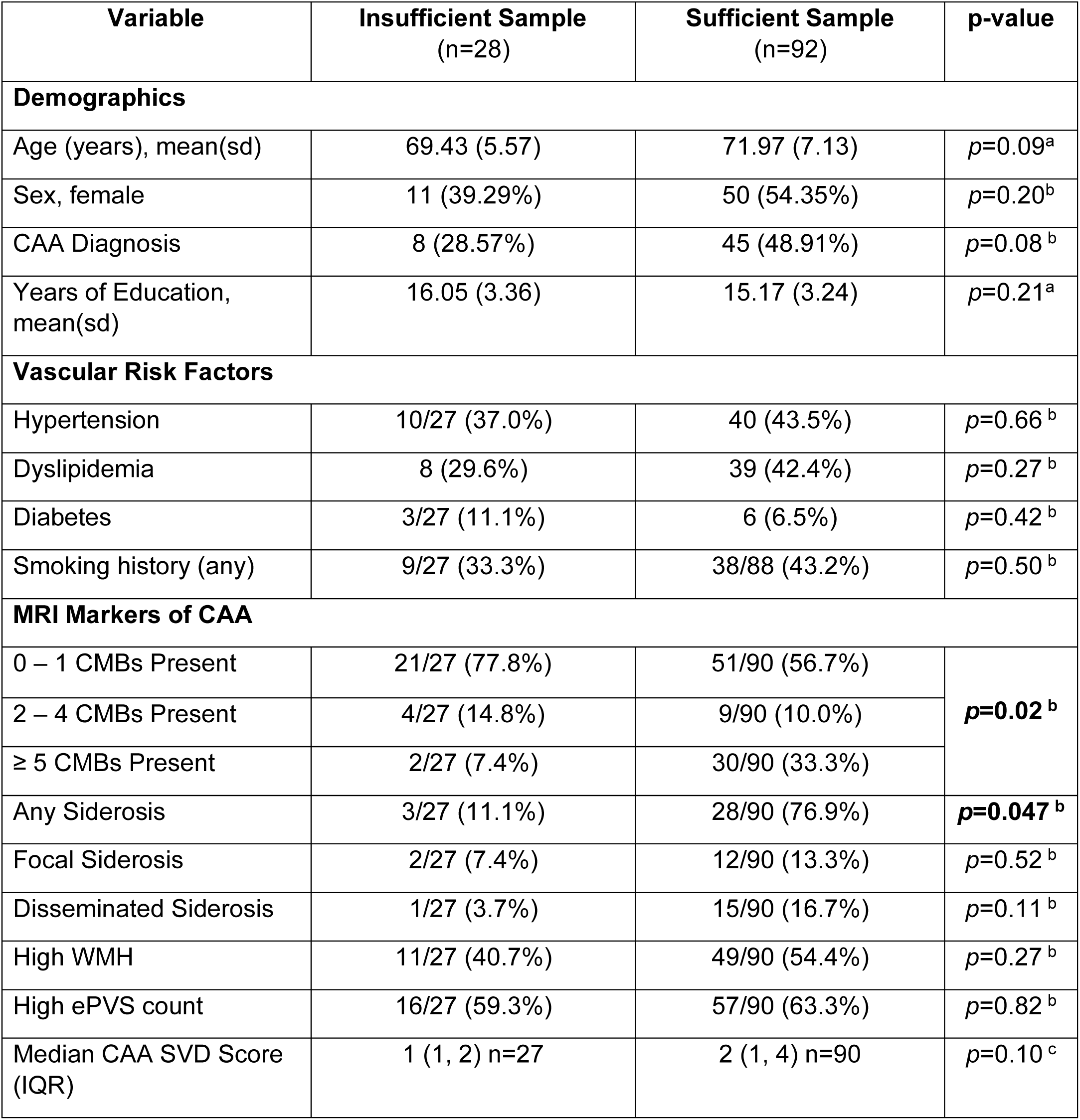
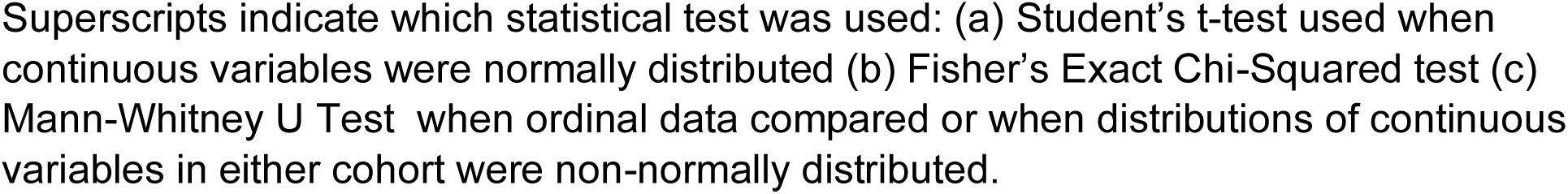
Demographic, Vascular Risk Factor, Cognitive and Neuroimaging Data comparing those with insufficient (n=28) sample to those with sufficient sample (n=92)

| Variable | Insufficient Sample<br>(n=28) | Sufficient Sample<br>(n=92) | p-value |
| --- | --- | --- | --- |
| **Demographics** |  |  |  |
| Age (years), mean(sd) | 69.43 (5.57) | 71.97 (7.13) | $p=0.09^a$ |
| Sex, female | 11 (39.29%) | 50 (54.35%) | $p=0.20^b$ |
| CAA Diagnosis | 8 (28.57%) | 45 (48.91%) | $p=0.08^b$ |
| Years of Education, mean(sd) | 16.05 (3.36) | 15.17 (3.24) | $p=0.21^a$ |
| **Vascular Risk Factors** |  |  |  |
| Hypertension | 10/27 (37.0%) | 40 (43.5%) | $p=0.66^b$ |
| Dyslipidemia | 8 (29.6%) | 39 (42.4%) | $p=0.27^b$ |
| Diabetes | 3/27 (11.1%) | 6 (6.5%) | $p=0.42^b$ |
| Smoking history (any) | 9/27 (33.3%) | 38/88 (43.2%) | $p=0.50^b$ |
| **MRI Markers of CAA** |  |  |  |
| 0 – 1 CMBs Present | 21/27 (77.8%) | 51/90 (56.7%) | $p=0.02^b$ |
| 2 – 4 CMBs Present | 4/27 (14.8%) | 9/90 (10.0%) |  |
| ≥ 5 CMBs Present | 2/27 (7.4%) | 30/90 (33.3%) |  |
| Any Siderosis | 3/27 (11.1%) | 28/90 (76.9%) | $p=0.047^b$ |
| Focal Siderosis | 2/27 (7.4%) | 12/90 (13.3%) | $p=0.52^b$ |
| Disseminated Siderosis | 1/27 (3.7%) | 15/90 (16.7%) | $p=0.11^b$ |
| High WMH | 11/27 (40.7%) | 49/90 (54.4%) | $p=0.27^b$ |
| High ePVS count | 16/27 (59.3%) | 57/90 (63.3%) | $p=0.82^b$ |
| Median CAA SVD Score (IQR) | 1 (1, 2) n=27 | 2 (1, 4) n=90 | $p=0.10^c$ |

Superscripts indicate which statistical test was used: (a) Student's t-test used when continuous variables were normally distributed (b) Fisher's Exact Chi-Squared test (c) Mann-Whitney U Test when ordinal data compared or when distributions of continuous variables in either cohort were non-normally distributed.

**Supplemental Table 5:** Comparison of demographic and vascular risk factors for n=11 that failed C2N quality control and n=81 who passed C2N quality control

| Variable | Passed C2N Quality Control (n=81) | Failed C2N Quality Control (n=11) | p-value |
| --- | --- | --- | --- |
| Age | 72.01 (7.18) | 71.69 (7.04) | $p=0.89^a$ |
| Sex, female | 45 (55.6%) | 5 (45.5%) | $p=0.54^b$ |
| CAA Diagnosis | 41 (50.6%) | 4 (36.4%) | $p=0.52^b$ |
| Years of Education | 15.07 (3.32) | 15.91 (2.66) | $p=0.42^a$ |
| Hypertension | 33 (40.7%) | 7 (63.6%) | $p=0.20^b$ |
| Dyslipidemia | 31 (38.3%) | 8 (72.7%) | **$p=0.048^b$** |
| Diabetes | 4 (4.9%) | 2 (18.2%) | $p=0.15^b$ |
| Smoking history (any) | 34/77 (44.2%) | 4 (36.4%) | $p=0.75^b$ |

Superscripts indicate which statistical test was used: (a) student's t-test used when continuous variables were normally distributed (b) Fisher's Exact Chi-Squared test (c) Mann-Whitney U Test when ordinal data compared or when distributions of continuous variables in either cohort were non-normally distributed

**Supplementary Table 6:** Linear regression models with age and sex adjusted standardized beta-coefficients for plasma biomarker values in CAA vs healthy controls

| Plasma Biomarker | Model | n | β-CAA | S.E | 95% CI: | p-value |
| --- | --- | --- | --- | --- | --- | --- |
| **Z-score $A\beta_{42/40}$ IPMS** | 1<sup>a</sup> | 81 | -0.67 | 0.21 | -1.09, -0.25 | $p=0.002$ |
| | 1<sup>b</sup> | 81 | -0.60 | 0.22 | -1.04, -0.15 | $p=0.009$ |
| | 1<sup>c</sup> | 81 | -0.39 | 0.22 | -0.83, 0.04 | $p=0.075$ |
| **Z-score $A\beta_{42/40}$ Simoa** | 2<sup>a</sup> | 75 | -0.77 | 0.21 | -1.20, -0.34 | $p=0.001$ |
| | 2<sup>b</sup> | 75 | -0.71 | 0.22 | -1.15, -0.27 | $p=0.002$ |
| | 2<sup>c</sup> | 75 | -0.69 | 0.23 | -1.15, -0.23 | $p=0.004$ |
| **Z-score log NfL pg/mL\*** | 3<sup>a</sup> | 75 | 1.06 | 0.20 | 0.67, 1.45 | $p<0.0001$ |
| | 3<sup>b</sup> | 75 | 0.86 | 0.19 | 0.49, 1.24 | $p<0.0001$ |
| | 3<sup>c</sup> | 75 | 0.93 | 0.19 | 0.54, 1.31 | $p<0.0001$ |
| **Z-score log p-tau-181 pg/mL\*** | 4<sup>a</sup> | 75 | 1.02 | 0.20 | 0.62, 1.42 | $p<0.0001$ |
| | 4<sup>b</sup> | 75 | 0.92 | 0.20 | 0.51, 1.33 | $p<0.0001$ |
| | 4<sup>c</sup> | 75 | 0.75 | 0.20 | 0.349, 1.14 | $p<0.0001$ |
| **Z-score log GFAP pg/mL\*** | 5<sup>a</sup> | 75 | 0.08 | 0.23 | -0.38, 0.54 | $p=0.735$ |
| | 5<sup>b</sup> | 75 | -0.19 | 0.24 | -0.50, 0.46 | $p=0.938$ |
| | 5<sup>c</sup> | 75 | 0.11 | 0.24 | -0.37, 0.60 | $p=0.641$ |

a. unadjusted model b. model solely adjusted for age c. model adjusted for age and sex
\* log transformation of the plasma biomarker concentration to improve normality of residuals and/or homoscedasticity in regression models, which was subsequently transformed to a Z score.

**Supplemental Table 7:** Proportion of Healthy Controls and those with CAA falling into age specific plasma concentration percentiles derived from n=900 in Cooper et al.^29^ from the ageing Canadian population for ages 3 – 79 years. Here the <5^th^ percentile was defined as anything less than the lower limit of the 95% CI of the predicted 5^th^ percentile for each age, while >95th percentile was defined as anything greater than the upper limit of the 95% CI of the predicted 95^th^ percentile for each age.

| **Biomarker** | **Group** | **5<sup>th</sup> – 95<sup>th</sup> percentile**<br>**Count(%)** | **<5<sup>th</sup> percentile**<br>**Count(%)** | **>95<sup>th</sup> percentile**<br>**Count(%)** | **Fisher's Exact Chi-Squared p-value** |
| --- | --- | --- | --- | --- | --- |
| Aβ<sub>42/40</sub> | healthy controls | 38/39 (97.44%) | 1/39 (2.56%) | 0/39 (0.00%) | *p*=0.014 |
|  | CAA | 28/36 (77.77%) | 7/36 (19.44%) | 1/36 (2.78%) |  |
| NfL | healthy controls | 35/39 (89.74%) | 1/39 (2.56%) | 3/39 (7.69%) | *p*<0.0001 |
|  | CAA | 19/36 (52.78%) | 0/36 (0.00%) | 17/36 (47.22%) |  |
| p-tau-181 | healthy controls | 33/39 (84.62%) | 5/39 (12.82%) | 1/39 (2.56%) | *p*=0.076 |
|  | CAA | 28/36 (77.78%) | 2/36 (5.56%) | 6/36 (16.67%) |  |
| GFAP | healthy controls | 33/39 (84.62%) | 5/39 (12.82%) | 1/39 (2.56%) | *p*=0.029 |
|  | CAA | 23/36 (69.44%) | 13/36 (36.11%) | 0/36 (0.00%) |  |

**Supplemental Table 8:** Number and Proportions of Healthy Controls and those with CAA categorized as having aberrantly low (<5^th^ percentile) Aβ_42/40_ or aberrantly high (>95^th^ percentile) elevations in p-tau-181, NfL, or GFAP (classified according to population reference intervals from Cooper et al.^29^)

| Biomarker | Group | Test Negative<br>$\geq 5\%$tile | Test Positive<br><5%tile | OR | 95% CI | p-value |
| --- | --- | --- | --- | --- | --- | --- |
| A$\beta_{42/40}$ | healthy controls | 38/39 (97.44%) | 1/39 (2.56%) | 9.17 | 1.07, 78.77 | $p=0.018$ |
|  | CAA | 29/36 (80.56%) | 7/36 (19.44%) |  |  |  |
| Biomarker | Group | Test Negative<br>$\leq 95\%$tile | Test Positive<br>>95%tile | OR | 95% CI | p-value |
| NfL | healthy controls | 36/39 (92.31%) | 3/39 (7.69%) | 10.74 | 2.79, 41.31 | $p=0.0001$ |
|  | CAA | 19/36 (52.78%) | 17/36 (47.22%) |  |  |  |
| p-tau-181 | healthy controls | 38/39 (97.44%) | 1/39 (2.56%) | 7.6 | 0.87, 66.59 | $p=0.036$ |
|  | CAA | 30/36 (83.33%) | 6/36 (16.67%) |  |  |  |
| GFAP | healthy controls | 37/39 (94.87%) | 2/39 (5.13%) | 0.0 | - | $p=0.17$ |
|  | CAA | 36/36 (100.00%) | 0/36 (0.00%) |  |  |  |

**Supplemental Table 9:**
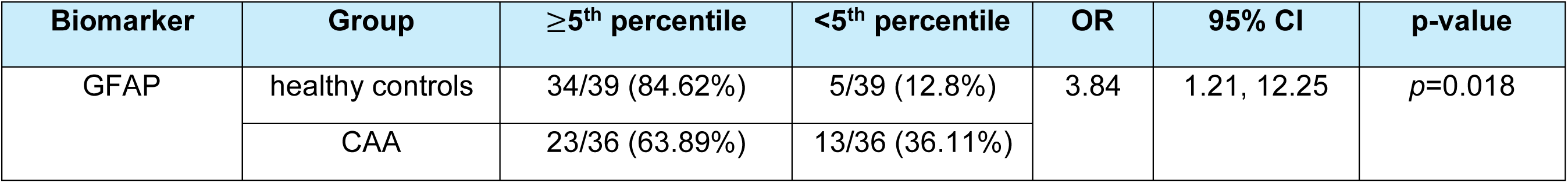
Number and Proportions of Healthy controls and those with CAA categorized with GFAP <5^th^ percentile (classified according to population reference intervals from Cooper et al.^29^)

| Biomarker | Group | ≥5<sup>th</sup> percentile | <5<sup>th</sup> percentile | OR | 95% CI | p-value |
| --- | --- | --- | --- | --- | --- | --- |
| GFAP | healthy controls | 34/39 (84.62%) | 5/39 (12.8%) | 3.84 | 1.21, 12.25 | *p*=0.018 |
|  | CAA | 23/36 (63.89%) | 13/36 (36.11%) |  |  |  |

**Supplemental Table 10:** Logistic Regression model beta-values (log-odds) of plasma-biomarkers in pg/mL and Youden’s Index defined univariate cut-points

| Model | Model Terms | β-coefficient | $\beta$ 95% CI | Cut point |
| --- | --- | --- | --- | --- |
| Model 1 | A$\beta_{42/40}$ IPMS | -81.22 | -136.73, -25.71 | 0.095 |
|  | *Constant Term* | 7.65 | 2.43, 12.87 |  |
| Model 2 | A$\beta_{42/40}$ Simoa | -79.01 | -28.38, -129.64 | 0.059 |
|  | *Constant Term* | 4.57 | 7.60, 1.54 |  |
| Model 3 | NfL Simoa (pg/mL) | 0.063 | 0.028, 0.097 | 23.22 pg/mL |
|  | *Constant Term* | -1.98 | -3.05, -0.92 |  |
| Model 4 | p-tau-181 Simoa (pg/mL) | 0.129 | 0.060, 0.197 | 32.24 pg/mL |
|  | *Constant Term* | -3.38 | -5.14, -1.62 |  |
| Model 5 | A$\beta_{42/40}$ IPMS | -11.62 | -86.93, 63.69 | 0.392 |
|  | NfL (pg/mL) | 0.087 | 0.028, 0.146 |  |
|  | p-tau-181 (pg/mL) | 0.067 | -0.019, 0.152 |  |
|  | *Constant Term* | -3.04 | -11.17, 5.09 |  |
| Model 6 | A$\beta_{42/40}$ Simoa | -86.33 | -142.24, -30.43 | 0.359 |
|  | NfL (pg/mL) | 0.049 | 0.009, 0.089 |  |
|  | p-tau-181 (pg/mL) | 0.094 | 0.022, 0.165 |  |
|  | *Constant Term* | 1.11 | -2.19, 4.41 |  |

## Supplementary Figures

**Supplementary Figure 1:**
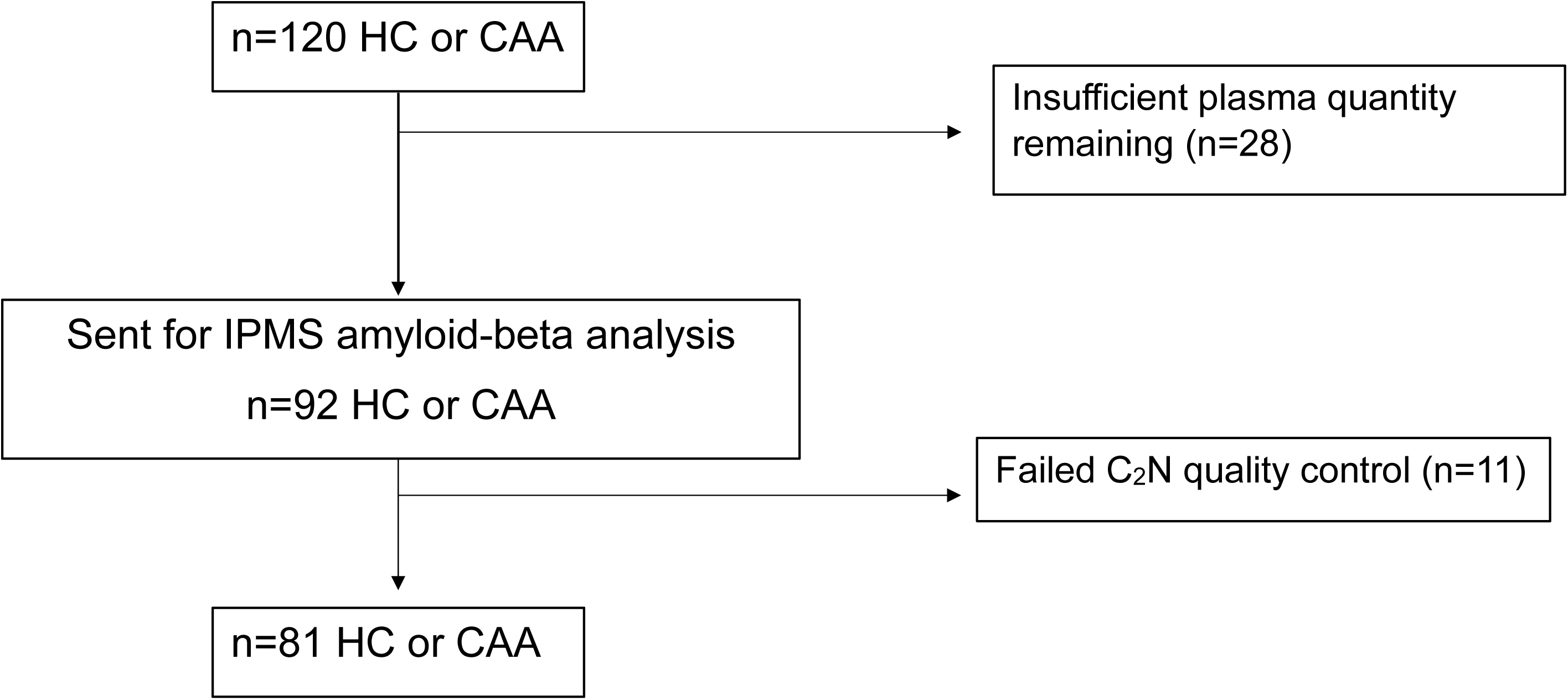
Patient flow diagram of consented participants in FAVR-II included in the IPMS amyloid-beta plasma biomarker analysis from C_2_N. Abbreviations: Healthy Controls (HC); Cerebral Amyloid angiopathy (CAA)

**Supplementary Figure 2:**
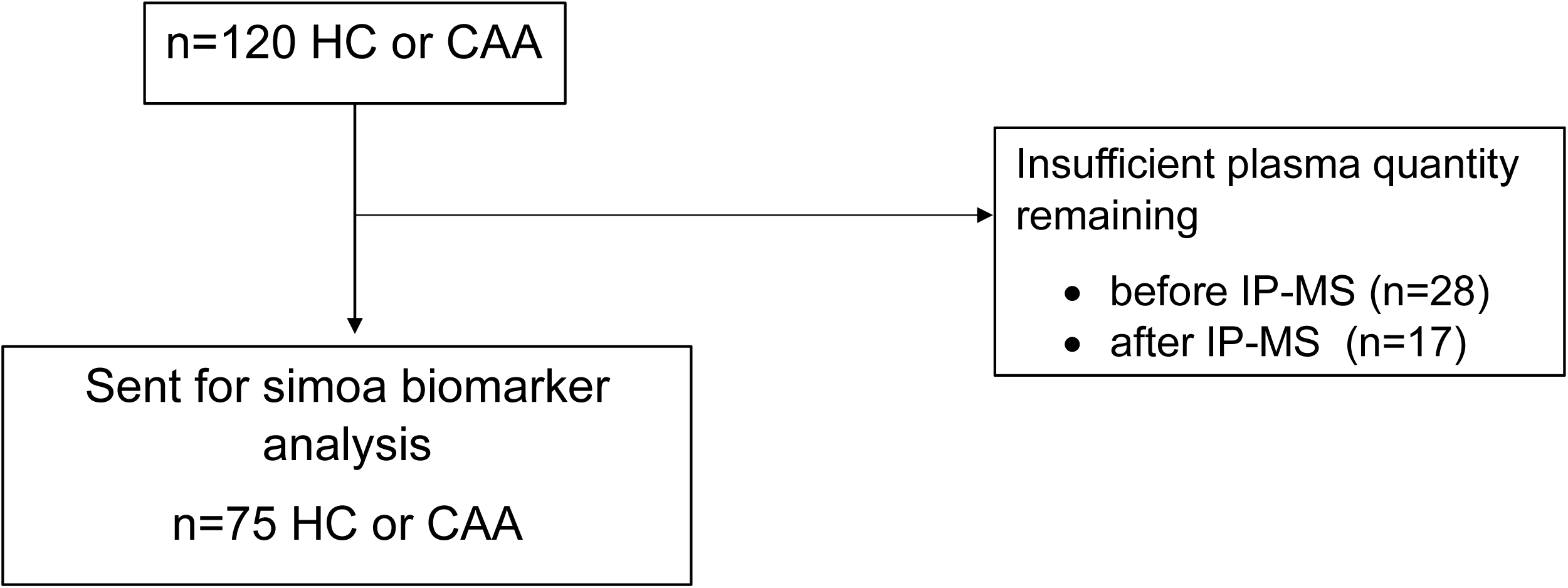
Patient flow diagram of consented participants included in the Simoa amyloid-beta (Quanterix Ltd), p-tau181, NfL and GFAP plasma biomarker analysis. Abbreviations: Healthy Controls (HC); Cerebral Amyloid angiopathy (CAA)

## REFERENCES

1. Quereshi AI, Mendelow AD, Hanley DF. Intracerebral haemorrhage. Lancet 2009;373:1632–1644.

2. Jolink WM KC, Brouwers PJ, Kappelle LJ, Vaartjes I. Time trends in incidence, case fatality, and mortality of intracerebral hemorrhage. Neurology 2015;85:1318–1324.

3. Sheth KN. Spontaneous Intracerebral Hemorrhage. N Engl J Med 2022;387:1589–1596.

4. Charidimou A, Boulouis G, Frosch MP, et al. The Boston criteria version 2.0 for cerebral amyloid angiopathy: a multicentre, retrospective, MRI–neuropathology diagnostic accuracy study. The Lancet Neurology 2022;21:714–725.

5. Boyle PA, Yu L, Leurgans SE, et al. Attributable risk of Alzheimer’s dementia attributed to age-related neuropathologies. Ann Neurol 2019;85:114–124.

6. Charidimou A, Imaizumi T, Moulin S, et al. Brain hemorrhage recurrence, small vessel disease type, and cerebral microbleeds. Neurology 2017;89:820–829.

7. Jakel L, De Kort AM, Klijn CJM, Schreuder F, Verbeek MM. Prevalence of cerebral amyloid angiopathy: A systematic review and meta-analysis. Alzheimers Dement 2022;18:10–28.

8. Moulin S, Labreuche J, Bombois S, et al. Dementia risk after spontaneous intracerebral haemorrhage: a prospective cohort study. Lancet Neurol 2016;15:820–829.

9. Hampel H, Elhage A, Cho M, Apostolova LG, Nicoll JAR, Atri A. Amyloid-related imaging abnormalities (ARIA): radiological, biological and clinical characteristics. Brain 2023;146:4414–4424.

10. Reish NJ, Jamshidi P, Stamm B, et al. Multiple Cerebral Hemorrhages in a Patient Receiving Lecanemab and Treated with t-PA for Stroke. N Engl J Med 2023;388:478–479.

11. Smith EE, Greenberg SM, Black SE. The impending era of beta-amyloid therapy: Clinical and research considerations for treating vascular contributions to neurodegeneration. Cerebral Circulation - Cognition and Behavior 2024;6.

12. van Dyck CH, Swanson CJ, Aisen P, et al. Lecanemab in Early Alzheimer’s Disease. N Engl J Med 2023;388:9–21.

13. Sims JR, Zimmer JA, Evans CD, et al. Donanemab in Early Symptomatic Alzheimer Disease: The TRAILBLAZER-ALZ 2 Randomized Clinical Trial. JAMA 2023;330:512–527.

14. Greenberg SM, Bacskai BJ, Hernandez-Guillamon M, Pruzin J, Sperling R, van Veluw SJ. Cerebral amyloid angiopathy and Alzheimer disease - one peptide, two pathways. Nat Rev Neurol 2020;16:30–42.

15. Koemans EA, Chhatwal JP, van Veluw SJ, et al. Progression of cerebral amyloid angiopathy: a pathophysiological framework. Lancet Neurol 2023;22:632–642.

16. Knopman DS, Amieva H, Petersen RC, et al. Alzheimer disease. Nat Rev Dis Primers 2021;7:1–47.

17. Switzer A, Charidimou A, McCarter SJ, et al. Boston Criteria v2.0 for Cerebral Amyloid Angiopathy Without Hemorrhage: An MRI-Neuropathologic Validation Study. Neurology 2024;102:e209386.

18. Zanon Zotin MC, Makkinejad N, Schneider JA, et al. Sensitivity and Specificity of the Boston Criteria Version 2.0 for the Diagnosis of Cerebral Amyloid Angiopathy in a Community-Based Sample. Neurology 2024;102:e207940.

19. Muir RT, Ismail Z, Black SE, Smith EE. Comparative methods for quantifying plasma biomarkers in Alzheimer’s disease: Implications for the next frontier in cerebral amyloid angiopathy diagnostics. Alzheimers Dement 2024;20:1436–1458.

20. Olsson B, Lautner R, Andreasson U, et al. CSF and blood biomarkers for the diagnosis of Alzheimer’s disease: a systematic review and meta-analysis. The Lancet Neurology 2016;15:673–684.

21. Sembill JA, Lusse C, Linnerbauer M, et al. Cerebrospinal fluid biomarkers for cerebral amyloid angiopathy. Brain Commun 2023;5:fcad159.

22. Beaudin AE, McCreary CR, Mazerolle EL, et al. Cerebrovascular Reactivity Across the Entire Brain in Cerebral Amyloid Angiopathy. Neurology 2022;98:e1716–e1728.

23. Charidimou A, Martinez-Ramirez S, Reijmer YD, et al. Total Magnetic Resonance Imaging Burden of Small Vessel Disease in Cerebral Amyloid Angiopathy: An Imaging-Pathologic Study of Concept Validation. JAMA Neurol 2016;73:994–1001.

24. Kirmess KM, Meyer MR, Holubasch MS, et al. The PrecivityAD test: Accurate and reliable LC-MS/MS assays for quantifying plasma amyloid beta 40 and 42 and apolipoprotein E proteotype for the assessment of brain amyloidosis. Clin Chim Acta 2021;519:267–275.

25. West T, Kirmess KM, Meyer MR, et al. A blood-based diagnostic test incorporating plasma Abeta42/40 ratio, ApoE proteotype, and age accurately identifies brain amyloid status: findings from a multi cohort validity analysis. Mol Neurodegener 2021;16:30.

26. Hu Y, Kirmess KM, Meyer MR, et al. Assessment of a Plasma Amyloid Probability Score to Estimate Amyloid Positron Emission Tomography Findings Among Adults With Cognitive Impairment. JAMA Netw Open 2022;5:e228392.

27. Fogelman I, West T, Braunstein JB, et al. Independent study demonstrates amyloid probability score accurately indicates amyloid pathology. Ann Clin Transl Neurol 2023;10:765–778.

28. Rissman RA, Langford O, Raman R, et al. Plasma Abeta42/Abeta40 and phospho-tau217 concentration ratios increase the accuracy of amyloid PET classification in preclinical Alzheimer’s disease. Alzheimers Dement 2024;20:1214–1224.

29. Cooper JG, Stukas S, Ghodsi M, et al. Age specific reference intervals for plasma biomarkers of neurodegeneration and neurotrauma in a Canadian population. Clin Biochem 2023;121–122:110680.

30. Shir D, Graff-Radford J, Hofrenning EI, et al. Association of plasma glial fibrillary acidic protein (GFAP) with neuroimaging of Alzheimer’s disease and vascular pathology. Alzheimers Dement (Amst) 2022;14:e12291.

31. Oeckl P, Halbgebauer S, Anderl-Straub S, et al. Glial Fibrillary Acidic Protein in Serum is Increased in Alzheimer’s Disease and Correlates with Cognitive Impairment. J Alzheimers Dis 2019;67:481–488.

32. Therriault J, Vermeiren M, Servaes S, et al. Association of Phosphorylated Tau Biomarkers With Amyloid Positron Emission Tomography vs Tau Positron Emission Tomography. JAMA Neurol 2023;80:188–199.

33. Williams S, Chalmers K, Wilcock GK, Love S. Relationship of neurofibrillary pathology to cerebral amyloid angiopathy in Alzheimer’s disease. Neuropathol Appl Neurobiol 2005;31:414–421.

34. Vidal R CM, Piccardo P, Farlow MR, Unverzagt FW, Méndez E, Jiménez-Huete A, Beavis R, Gallo G, Gomez-Tortosa E, Ghiso J, Hyman BT, Frangione B, Ghetti B. Senile dementia associated with amyloid beta protein angiopathy and tau perivascular pathology but not neuritic plaques in patients homozygous for the APOE-epsilon4 allele. Acta Neuropathol 2000;100:1–12.

35. Grangeon L, Paquet C, Guey S, et al. Cerebrospinal Fluid Profile of Tau, Phosphorylated Tau, Abeta42, and Abeta40 in Probable Cerebral Amyloid Angiopathy. J Alzheimers Dis 2022;87:791-802.

36. Boyle PA, Wang T, Yu L, et al. To what degree is late life cognitive decline driven by age-related neuropathologies? Brain 2021;144:2166–2175.

37. Rabin JS, Nichols E, La Joie R, et al. Cerebral amyloid angiopathy interacts with neuritic amyloid plaques to promote tau and cognitive decline. Brain 2022;145:2823–2833.

38. Benedet AL, Mila-Aloma M, Vrillon A, et al. Differences Between Plasma and Cerebrospinal Fluid Glial Fibrillary Acidic Protein Levels Across the Alzheimer Disease Continuum. JAMA Neurol 2021;78:1471–1483.

39. Stocker H, Beyer L, Perna L, et al. Association of plasma biomarkers, p-tau181, glial fibrillary acidic protein, and neurofilament light, with intermediate and long-term clinical Alzheimer’s disease risk: Results from a prospective cohort followed over 17 years. Alzheimers Dement 2022;19:25-35.

40. Benedet AL, Smirnov DS, Ashton NJ, et al. Neuropathological validation of plasma GFAP. Alzheimer’s & Dementia 2022;18.

41. Charidimou A, Friedrich JO, Greenberg SM, Viswanathan A. Core cerebrospinal fluid biomarker profile in cerebral amyloid angiopathy: A meta-analysis. Neurology 2018;90:e754–e762.

42. Sembill JA, Lusse C, Linnerbauer M, et al. Cerebrospinal fluid biomarkers for cerebral amyloid angiopathy. Brain Commun 2023.

43. Theodorou A, Tsantzali I, Stefanou MI, et al. CSF and plasma biomarkers in cerebral amyloid angiopathy: A single-center study and a systematic review/meta-analysis. Eur Stroke J 2024:23969873241260538.

44. Cheng X, Su Y, Wang Q, et al. Neurofilament light chain predicts risk of recurrence in cerebral amyloid angiopathy-related intracerebral hemorrhage. Aging 2020;12:23727–23738.

45. De Kort AM, Kuiperij HB, Jakel L, et al. Plasma amyloid beta 42 is a biomarker for patients with hereditary, but not sporadic, cerebral amyloid angiopathy. Alzheimers Res Ther 2023;15:102.

46. Chatterjee P, Tegg M, Pedrini S, et al. Plasma Amyloid-Beta Levels in a Pre-Symptomatic Dutch-Type Hereditary Cerebral Amyloid Angiopathy Pedigree: A Cross-Sectional and Longitudinal Investigation. Int J Mol Sci 2021;22:2931.

47. Greenberg SM, Cho HS, O’Donnell HC, et al. Plasma beta-amyloid peptide, transforming growth factor-beta 1, and risk for cerebral amyloid angiopathy. Ann N Y Acad Sci 2000;903:144–149.

48. Piccarducci R, Caselli MC, Zappelli E, et al. The Role of Amyloid-beta, Tau, and alpha-Synuclein Proteins as Putative Blood Biomarkers in Patients with Cerebral Amyloid Angiopathy. J Alzheimers Dis 2022;89:1039–1049.

49. Hernandez-Guillamon M, Delgado P, Penalba A, et al. Plasma beta-amyloid levels in cerebral amyloid angiopathy-associated hemorrhagic stroke. Neurodegener Dis 2012;10:320–323.

50. Charidimou A, Boulouis G, Gurol ME, et al. Emerging concepts in sporadic cerebral amyloid angiopathy. Brain 2017;140:1829–1850.

51. Quanterix. Simoa p-tau 181 Advantage V2.1 Assay (reference TECH-0153). 2022. Available at: https://portal.quanterix.com/folder-viewer/Assay%20Technical%20Notes/Bead%20Assays.

